# Prediction models for longitudinal trajectories of depression and anxiety: a systematic review

**DOI:** 10.1101/2025.10.09.25337650

**Authors:** Sophie J Fairweather, Holly Fraser, Natalie Lam, Simon Gilbody, Lewis W Paton, Hannah J Jones, Golam M Khandaker

## Abstract

**Background:** Prediction of atypical health trajectories may enable early intervention. We systematically reviewed the existing literature on models for predicting longitudinal depression and/or anxiety trajectories.

**Methods:** MEDLINE, Embase and APA PsycINFO were searched (from inception to 31-Jan-2025). We included population-based studies of children and adults (aged 3-65 years). We extracted data using the CHecklist for critical Appraisal and data extraction for systematic Reviews of prediction Modelling Studies (CHARMS). Risk of bias was assessed using the Prediction model Risk Of Bias ASsessment Tool (PROBAST-AI) tool.

**Results:** Seven of the nine included studies were in adult populations with a diagnosis of depression or anxiety at baseline; two focused on child and adolescent populations. Only one study included anxiety trajectories. Identified trajectories typically comprised three to four groups including: chronic/persistent-high, stable-low, increasing/worsening, and improved/remitted groups. A range of supervised predictive modelling methods were used. The number of final predictors included in models ranged from three to 152. Family and own/personal psychiatric history were the most common predictors but were not always important for model performance. Models including a large number of predictors did not always perform better. Overall risk of bias was high across all studies. No studies were externally validated and no studies assessed the clinical utility of models.

**Conclusion:** This review highlights a need for robust, validated models that can forecast future risk of persistent or worsening anxiety and depression, especially in young people where early intervention is possible.

**PROSPERO; ID:** CRD42024628610

## 1. Background

Despite extensive research into individual risk and prognostic factors for depression and anxiety (Buckman et al., 2018; Burcusa and Iacono, 2007; Musliner et al., 2016; Shore et al., 2018; Struijs et al., 2021), we lack prediction tools for forecasting future outcomes at the individual level (Meehan et al., 2022). Approximately half of depression cases first emerge before age 30 and nearly 40% of anxiety cases emerge before age 14 (Solmi et al., 2022). Early prediction of individuals at high-risk of persistent or worsening symptom trajectories may offer richer insights than predicting outcomes like diagnosis or relapse at a single timepoint. Prediction of trajectories could transform how we target early intervention, before symptoms escalate, to reduce the long-term impact of anxiety and depression.

Depression and anxiety are common mental disorders. In recent decades, the incidence of anxiety and depression in young people has risen sharply in developed countries (Bie et al., 2024; Dongjun et al., 2025; Hua et al., 2024; Liu et al., 2024). Early onset is associated with continued symptoms into adulthood (Portogallo et al., 2024), higher physical health risks(Blasco et al., 2020; Inoue et al., 2020; Naicker et al., 2013) and NEET (“Not in education, employment or training”) status (Crowley et al., 2023; Rahmani and Groot, 2023).

Prediction research in mental health so far has mostly focused on predicting the course of illness in adults with depression (King et al., 2013; Meehan et al., 2022; Moriarty et al., 2022; Senior et al., 2021; Todd et al., 2025). Outcomes in models identified by previous systematic reviews include relapse and recurrence measured at a single timepoint (Allesøe et al., 2023; Kieling et al., 2021; King et al., 2013; Meehan et al., 2022; Moriarty et al., 2022, 2024; Senior et al., 2021; Todd et al., 2025). While being able to forecast these outcomes is useful, traditional definitions of concepts like “relapse” and “remission” oversimplify the fluctuating nature of depression/anxiety symptoms and require arbitrary timeframes (Buckman et al., 2018). Conversely, studying symptom trajectories captures onset and heterogeneity in illness course. Trajectories are useful because different people may arrive at the same destination (e.g., severely depressed) by different routes (e.g., sudden onset or gradual development). Early prediction could enable more personalised intervention to reduce the long-term impact of anxiety and depression.

Few models have been developed for predicting onset of depression and anxiety in young people (Meehan et al., 2022; Senior et al., 2021). In child/adolescent mental health more broadly, outcomes of prediction models have included diagnoses of depression and neurodevelopmental conditions (Senior et al., 2021). Generally, models predicting anxiety are scarce and this is notable in young people given the early onset of anxiety (Solmi et al., 2022). A promising model for predicting depression *onset* in adolescence has been developed and validated in international cohorts with moderate predictive performance (C-statistics: 0.59-0.78, 95%CIs 0.53-0.83) (Kieling et al., 2021; Piccin et al., 2025; Rocha et al., 2021). This suggests predicting onset in adolescence is feasible. However, the model was not developed to predict trajectories, or anxiety outcomes.

Currently, there are no systematic reviews of multivariable prediction models for longitudinal symptom trajectories of depression and/or anxiety. Existing studies suggest that risk factors including female gender and low socioeconomic status are *associated with* persistently high and increasing symptom trajectories in *single* variable prediction models (Musliner et al., 2016; Shore et al., 2018). How these factors perform in multivariable prediction models remains unknown.

We aimed to systematically identify and examine multivariable prediction models for longitudinal trajectories of anxiety and/or depression. Our objectives were to:

- Systematically identify models for predicting longitudinal trends/patterns/trajectories in depression and/or anxiety in general population samples.
- Review model characteristics, including: predictors used, development and validation methods, and predictive performance.
- Assess quality and risk of bias of the included studies.
- Make recommendations for future development of prediction models for depression/anxiety trajectories.

## 2. Methods

This study was prospectively registered (PROSPERO ID:CRD42024628610). We followed the Preferred Reporting Items for Systematic Reviews and Meta-Analyses (PRISMA) guidelines (Moher et al., 2015; Page et al., 2021) (Supplementary tables s1 and s2: PRISMA checklists). Protocol deviations were minor (see supplement).

### Eligibility criteria

We pre-specified the following key eligibility criteria:

- **Population:** General population, adults and children aged 3-65 years.
- **Index model:** Any multivariable prediction model for predicting below outcome.
- **Comparator:** Single model or comparison of performance across multiple models.
- **Outcome (for which model is validated to predict):** Longitudinal trajectories of anxiety and/or depression diagnosis/symptom(s) (using any measurement scale/instrument e.g., self or proxy reports, questionnaire scores, diagnoses from health records). We report performance measures of the included prediction models for predicting trajectories.
- **Timeframe (prediction horizon):** Any time horizon.
- **Setting:** Population based (including primary care).

See supplementary information for extended eligibility criteria.

Our primary interest was prediction models for predicting depression/anxiety trajectories. We included any study aimed at developing, validating, and/or adjusting/extending multivariable prediction/prognostic model(s) for making predictions in individuals. We excluded studies of single prediction factors, and model development studies without an internal validation step. We used the term longitudinal trends/patterns/trajectories to encompass all studies/methods using repeated anxiety and/or depression measures to quantitatively model change over time. We excluded studies measuring only change between two timepoints as such studies show the *direction* or *magnitude* of one change rather than a longitudinal pattern.

Eligible outcomes had a trajectory start point between ages 3 and 65 (inclusive). Studies *exclusively* modelling trajectories of other psychiatric conditions, and studies conducted in specialist settings/populations, were excluded. We were not directly interested in the statistical development of trajectories themselves. Further details in supplement.

### Search strategy

We searched OVID MEDLINE, Embase, and APA PsycINFO from inception to 31-January-2025. Studies were limited to English language and full-text records. No date restrictions were applied. Search terms were based on four concepts: “prediction/prognostic model” AND “trajectories” AND “anxiety” OR “depression”. We defined the full list of search terms based on high-sensitivity published filters (Glanville et al., 2022) for prognosis reviews to ensure we captured all relevant studies (supplementary information: full search strategy). Additionally, we hand searched related systematic reviews and searched grey literature for eligible studies (medRvix, preprints.org and psyArXiv).

### Study selection

Two reviewers independently screened titles/abstracts of articles in Rayyan (Ouzzani et al., 2016) against the pre-specified eligibility criteria. Full-text reports of potentially eligible studies were obtained, and at least two reviewers assessed the eligibility. Disagreements were resolved by discussion between reviewers (SJF, NL, HF, HJJ); a third reviewer (GK, LP) was consulted when necessary. Reason for exclusion at full-text stage is summarised in Figure 1 (PRISMA diagram) and reported in full in supplementary information.

**Figure 1.**
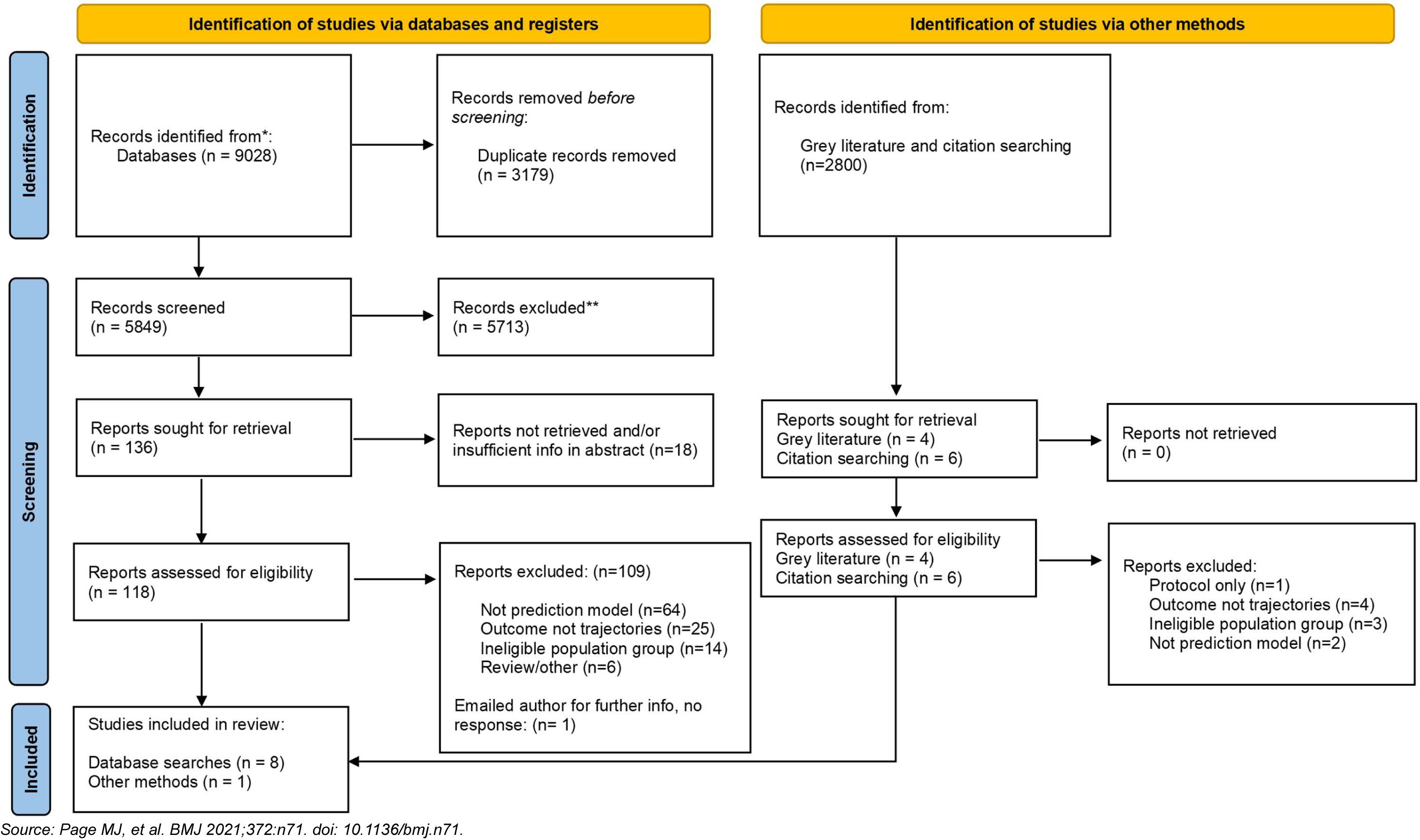
PRISMA diagram: study screening, inclusion and exclusion.

### Data extraction and risk of bias assessment

We extracted data using the CHecklist for critical Appraisal and data extraction for systematic Reviews of prediction Modelling Studies (CHARMS) (Fernandez-Felix et al., 2023). We used the Prediction model Risk of Bias ASsessment Tool (PROBAST-AI, 2025) (Moons et al., 2025) to assess risk of bias (RoB) for the best performing model from each study. Data extraction and RoB assessments were performed by one reviewer (SF) and checked by a second reviewer (HJJ, HF).

We extracted key study and model characteristics for each prediction model. We report performance statistics for the best performing model from each paper in the main text (supplementary sTables 6 and 7: data extracted from additional models). We calculated events-per-variable (EPV) for multinomial and binomial models (formulae and calculations reported in supplementary methods, sTables 3-6).

Studies were judged as having high, low or unclear RoB across four domains: participants, predictors, outcome and analysis. For participant/predictor/outcome domains an “applicability” rating was assigned referring to alignment between the included study vs. the review question.

We report findings as a narrative synthesis. Meta-analysis was unsuitable due to heterogeneity and a lack of validation studies of the same index model (Damen et al., 2023). We present the frequency and importance of key predictor types across studies in a bar chart.

## 3. Results

Nine studies were eligible and included (Figure 1), which together reported 45 unique prediction models (Dinga et al., 2018; Gorham et al., 2022; Kessler et al., 2016; Schultebraucks et al., 2021; Teutenberg et al., 2025; van Loo et al., 2014; Wardenaar et al., 2021, 2014; Xiang et al., 2022). Performance was reported for 37 of the 45 models.

### Study characteristics

Characteristics of included studies are shown in Table 1. We identified eight model development studies and one external validation study (Kessler et al., 2016), although the validation model used different predictors to the two model development studies (van Loo et al., 2014; Wardenaar et al., 2014).

**Table 1.** Characteristics of included studies.

| Study ID | Study type<br>(model<br>development<br>vs validation) | Parent cohort / data source for the studies included in the review |  |  |  | Baseline sample characteristics of studies included in the review |  |  |  |
| --- | --- | --- | --- | --- | --- | --- | --- | --- | --- |
|  |  | Data source, type, and location | Enrolment period | Study setting | Age range of cohort (years) | Total sample at baseline | Mean sample age (years) | Proportion female (%) | Anxiety or depression status at baseline |
| Teutenberg 2025 (Teutenberg et al., 2025) | Development | MACS; Prospective cohort; Germany | 2014 - 2018 | Population, primary and secondary care | 18-65 | 571 | 36.7 (±13.2) | 67% | MDD at study baseline |
| Gorham 2022 (Gorham et al., 2022) | Development | NIMH CAT-D Prospective cohort; USA | 2017 | Population, primary and secondary care | 11-17 | 92 | 15.8 (±1.3) | 72% | Mixed (healthy and depressed) |
| Xiang 2022 (Xiang et al., 2022) | Development | ABCD study; Prospective cohort; USA | 2016-2018 | Population based | 9-10 | 4962 | Not given | 49% | Healthy |
| Schultebrasucks 2021 (Schultebrasucks et al., 2021) | Development | Health and Retirement study; Prospective cohort; USA | 1992 | Population based | 50+ | 2071 | 55.9 (±8.5) | 64% | Healthy |
| Wardenaar 2021 (Wardenaar et al., 2021) | Development | NESDA; Prospective cohort; Netherlands | 2004-2006 | Population, primary and secondary care | 18-65 | 1693 | 41.3 (±12.4) | 67% | MDD, Dysthymia, GAD or other anxiety disorder in past 6m |
| Dinga 2018 (Dinga et al., 2018) | Development | NESDA; Prospective cohort; Netherlands | 2004-2006 | Population, primary and secondary care | 18-65 | 804 | 41.9 (±12.2) | 65% | MDD or dysthymia |
| Kessler 2016 (Kessler et al., 2016) | Validation (of Van Loo and Wardenaar 2014 papers) | US National Comorbidity survey 1 & 2; longitudinal follow up of sub-sample from cross-sectional national survey; USA | 1990-1992 | Population based | 15-54 | 1056 | Not given | 50% | MDD at study baseline |
| Van Loo 2014 (van Loo et al., 2014) | Development | WMH Surveys; Retrospective cohort; Multi-country (n=16) | 2001-2009 | Population based | 18+ | 8261 | Not given | Not given | Lifetime MDD |
| Wardenaar 2014 (Wardenaar et al., 2014) | Development | WMH Surveys; Retrospective cohort; Multi-country (n=16) | 2001-2009 | Population based | 18+ | 8261 | Not given | Not given | Lifetime MDD |

Footnotes: ABCD=Adolescent Brain Cognitive Development study, USA=United States of America, NESDA=Netherlands Study of Depression and Anxiety, MACS=Marburg- Münster Affective Disorders Cohort Study, NL=Netherlands, WMH=world mental health, NIMH CAT-D=National Institute of Mental Health Characterization and Treatment of Adolescent Depression, MDD=major depressive disorder, GAD=generalized anxiety disorder

Seven studies were in adult populations (Dinga et al., 2018; Kessler et al., 2016; Schultebraucks et al., 2021; Teutenberg et al., 2025; van Loo et al., 2014; Wardenaar et al., 2021, 2014). Six of these were in adults with a diagnosis of depression at baseline (Dinga et al., 2018; Kessler et al., 2016; Teutenberg et al., 2025; van Loo et al., 2014; Wardenaar et al., 2021, 2014). Two studies were in child (9-10 years) and adolescent populations (11-17 years) (Gorham et al., 2022; Xiang et al., 2022); one in participants without baseline depression (Xiang et al., 2022), one in a mixed population of healthy adolescents and adolescents with subthreshold illness or a depression diagnosis (Gorham et al., 2022). Only one study included anxiety trajectories alongside depression trajectories (Wardenaar et al., 2021); no studies solely studied anxiety trajectories. Most study samples had more females than males (>60% in five studies).

Seven studies used data from prospective cohorts (Dinga et al., 2018; Gorham et al., 2022; Kessler et al., 2016; Schultebraucks et al., 2021; Teutenberg et al., 2025; Wardenaar et al., 2021; Xiang et al., 2022). Two studies (van Loo et al., 2014; Wardenaar et al., 2014) collected depression outcome data retrospectively for multiple timepoints which were then used to characterise depression outcomes over time. Four studies were conducted using cohorts from the USA (Gorham et al., 2022; Kessler et al., 2016; Schultebraucks et al., 2021; Xiang et al., 2022) and two from the Netherlands (Dinga et al., 2018; Wardenaar et al., 2021) (both using data from Netherlands Study of Depression and Anxiety [NESDA]).

### Trajectory modelling methods and trajectories identified

Studies differed in how they defined and analysed trajectories. Six studies used recognised, class-based trajectory modelling approaches to delineate trajectories e.g., latent class growth analysis (LCGA). One study calculated the number of weeks spent in a depressive episode from baseline to one year follow up (Gorham et al., 2022). The remaining two studies quantified depression over time as proportion of years since age of onset where subject had MDD. This was sub-categorised as: 1) “chronic MDD”: proportion of years with symptoms lasting most days of the year; and 2) “persistent MDD”: proportion of years with MDD episodes lasting two or more weeks (van Loo et al., 2014; Wardenaar et al., 2014) (Table 2).

**Table 2.** Sub-groups and trajectories identified as the outcomes for prediction model in the included studies.

| Study ID | Outcome measure | Trajectory modelling method, number of datapoints, and length of follow-up | Trajectories/sub-groups identified and sample size (n, %) |
| --- | --- | --- | --- |
| Teutenberg 2025 | MDD (SCID-I) | Model-based clustering approach and latent profile analysis<br>24 timepoints; reported retrospectively by life charting method for each month over 2 year interval | Remitted (n=178, 65%)<br>Dysthymic (n=30, 11%)<br>Moderate (n=47, 17%)<br>Severe (n=18, 7%) |
| Gorham 2022 | Depression (K-SADS-PL DSM 5 depression screener and supplement) | Number of weeks spent in depressive episode from baseline to 1 year follow up | n/a - mean number of weeks for sample not given |
| Xiang 2022 | Depressive symptoms (CBCL) | LCGA<br>3 timepoints over 2 year follow up | Persistently low (n=3724, 75%)<br>Decreasing (n=433, 9%)<br>Increasing (n=536, 11%)<br>Persistently high (n=269, 5%) |
| Schultebrasucks 2021 | Depression after single major life stressor event (CES-D) | LGMM<br>3 timepoints over 2 year follow up | Resilience (n=1638, 79%)<br>Recovery (n=160, 8%)<br>Emerging depression (n=159, 8%)<br>Pre-existing and chronic depression (n=114, 5%) |
| Wardenaar, 2021 | Depression and Anxiety (IDS-SR, BAI) | LCGA<br>5 timepoints over 9 year follow up | Depression, chronic (n=1078, 64%)<br>Depression, partial recovery (n=502, 30%)<br>Depression, full recovery (n=112, 6%)<br>Anxiety, full recovery (n=236, 14%)<br>Anxiety, partial recovery (n=1306, 77%)<br>Anxiety, increasing severity (n=151, 9%) |
| Dinga 2018 | MDD diagnosis (CIDI) | LCGA<br>24 timepoint over 2 year follow up | Remitted (n=356, 44%)<br>Improved (n=273, 34%)<br>Chronic (n=175, 22%) |
| Kessler 2016 | MDD diagnosis (CIDI) | Proportion of years since age of onset where subject had MDD over 10-12 year follow up | Persistent (years with episode 2+ weeks) (n=not reported)*<br>Chronic (lasting most days in year) (n=176)<br>*Mean (se) number of years in episode = 2.0 (0.2) |
| Van Loo 2014 | MDD (CIDI) | Proportion of years since age of onset where subject had MDD over 10-12 year follow up | Persistent (yrs with episode 2+ wks) (n=2869)*<br>Chronic (lasting most days in year) (n=3958)<br>Mean proportion of follow up with persistent depression=25.8%<br>Mean proportion of follow up with chronic depression=9.5% |
| Wardenaar 2014 | MDD (CIDI) | Proportion of years since age of onset where subject had MDD over 10-12 year follow up | Persistent (years with episode 2+ weeks) (n=2869)*<br>Chronic (lasting most days in year) (n=3958)* |

\*Percentage not reported as groups are not discrete and pay overlap. Footnotes: CBCL=child behaviour checklist, IDS-SR=inventory of depressive symptomatology: self-report, MDD=major depressive disorder, CIDI=composite international diagnostic interview, K-SADS-PL DSM-5=Kiddie schedule for Affective disorders and Schizophrenia, CES-D=centre for epidemiologic studies depression scale, SCID= Structured Clinical Interview for DSM-IV disorders, LCGA=latent class growth analyses, LGMM=latent growth mixture modelling

The number of identified trajectories ranged from three to four, typically including chronic/persistent-high, stable-low, increasing/worsening, and improved/remitted/decreasing groups. Different terminology was used across studies to describe similar patterns. The time-horizon of trajectories ranged from 1-12 years. The number of repeated data points used to model trajectories ranged between 3-24. Studies with longer time-horizons did not necessarily have more timepoints. For example, Teutenberg et al., 2025 (Teutenberg et al., 2025) modelled trajectories with 24 timepoints over two years, whereas Wardenaar et al., (2021) (Wardenaar et al., 2021) used five timepoints over nine year

### Prediction models characteristics and performance

Key model characteristics and performance statistics for the best performing model from each included study are summarised in Table 3. Further characteristics are detailed in the supplement along with performance of additional models (sTable 7).

**Table 3.** Key characteristics of the best performing prediction models from each study.

| Study ID | Model name | Modelling method and outcome type | Predictors used in final model, No. | Main performance statistics | Other performance measures |
| --- | --- | --- | --- | --- | --- |
| Teutenberg 2025<br>(Teutenberg et al., 2025) | Explorative model | One-vs-all classification using random forest (categorical) | 20 | **Discrimination:**<br>Remitted; AUC = 74.38 (0.26)<br>Dysthymic; AUC = 51.73 (3.45)<br>Moderate; AUC = 63.59 (3.71)<br>Severe; AUC = 67.63 (0.93)<br>Note: performance in internal validation sample | **Remitted;** BACC= 67.14 (1.21), sens= 56.86 (3.51), spec= 77.42 (3.26), PPV= 83.85 (1.34), NPV = 46.69 (1.35)<br>**Dysthymic;** BACC= 52.72 (1.91), sens= 24.24 (5.42), spec= 81.20 (3.82), PPV= 14.60 (2.10), NPV= 89.08 (0.50)<br>**Moderate;** BACC= 52.35 (2.71), sens=16.17 (8.14), spec = 88.52 (4.02), PPV =20.87 (5.72), NPV=84.22 (0.87)<br>**Severe;** BACC= 66.33 (2.96), sens=53.85 (9.73), spec=78.82(4.44), PPV=10.97(0.99), NPV=97.29 (0.44) |
| Xiang 2022 (Xiang et al., 2022) | GBM Model | Multinomial classification (categorical) | 24 | **Discrimination:** Micro-average AUC = 0.90 (no CIs given)<br>Macro-average AUC = 0.77 (no CIs given) | **Macro-average:**<br>Accuracy = 0.87<br>Specificity = 0.82<br>F1 score = 0.45<br>Precision = 0.45<br>Recall = 0.45 |
| Gorham 2022 (Gorham et al., 2022) | MFQ+FH+CASE | Regression (no further information) (continuous) | 10 | RMSE (in weeks): 15.3 (99.9% CI not given for this model) | None reported |
| Schultebracks 2021<br>(Schultebrack s et al., 2021) | Neural network: | Multinomial classification and individual discrimination (categorical) | 21 | **Discrimination:**<br>**Multinomial:**<br>Macro-average AUC = 0.86 (95% CI, 0.85-0.87)<br>Micro-average AUC = 0.88 (95% CI, 0.87-0.89)<br>**Per trajectory:**<br>Recovery AUC = 0.87<br>Emerging AUC = 0.88<br>Chronic AUC = 0.93<br>Resilience AUC = 0.75 | Average precision, 0.79; average recall, 0.60; average specificity, 0.82; average F1, 0.64; average geometric mean, 0.70; and average index balanced accuracy, 0.48 |
| Wardenaar 2021 (Wardenaar et al., 2021) | Super Learner Model | Probabilistic multi-classification (continuous) | 152 | **Depression:**<br>Chronic course MSE = 0.131(±0.003), ppred o/e=0.81<br>Partial-recovery MSE = 0.114(±0.003), ppred o/e=0.82<br>Full-recovery MSE = 0.049(±0.004), ppred o/e=0.60<br>**Anxiety:**<br>Full recovery MSE = 0.095(±0.005), ppred o/e=0.53<br>Partial-recovery MSE = 0.116(±0.004), ppred o/e=0.88<br>Increasing severity MSE = 0.057 (±0.004), ppred o/e=0.73 | None reported |
| Dinga 2018 (Dinga et al., 2018) | Full model | Multinomial classification and one-vs-all discrimination using: penalized (elastic-net) logistic regression | 81 | **Discrimination:**<br>Remitted AUC = 0.69<br>Improved AUC = 0.62<br>Chronic AUC = 0.66 | **One-vs-all class:**<br>Remitted; Balanced accuracy = 66%<br>Improved; Balanced accuracy = 60%<br>Chronic; Balanced accuracy = 61%<br>**Multinomial prediction:**<br>Remitted; sensitivity = 59%<br>Improved; sensitivity = 37%<br>Chronic; sensitivity = 47% |
| Kessler 2016 (Kessler et al., 2016) | Ensemble regression tree | Ensemble regression trees, logistic regression (continuous) | Between 9-13 | **Discrimination:**<br>Persistent, AUC = 0.71 (no CIs given)<br>Chronic, AUC = 0.63 (no CIs given) | **Sensitivity:**<br>Highest 20% of persistence: 38.1(4.2)<br>Lowest 20% of persistence: 5.6 (1.8)<br>Highest 20% of chronic: 34.6 (7.3)<br>Lowest 20% of chronic: 15.9 (5.8) |
| Van Loo 2014 (van Loo et al., 2014) | Three-cluster | GLM with Lasso penalised regression (continuous) | –Unclear – at least 29. | **Discrimination:**<br>Persistent, AUC = 0.64 (no CIs given)<br>Chronic, AUC = 0.61 (no CIs given)<br>*No performance statistics of GLM multivariate prediction model reported* | None reported |
| Wardenaar 2014 (Wardenaar et al., 2014) | Three-cluster | GLM with Lasso penalised regression (continuous) | Unclear.- at least 41 | **Discrimination:**<br>Persistent, AUC = 0.68 (no CIs given)<br>Chronic, AUC = 0.62 (no CIs given)<br>*No performance statistics of GLM multivariate prediction model reported (?)* | None reported |

**Abbreviations:** GBM=gradient boosting machine, N=number, AUC=area-under-curve, MSE=mean standard error, RMSE=root mean squared error, GLM=generalized linear model, ppred o/e= Spearman correlation between SL-predicted and observed scores, BACC=balanced accuracy, sens=sensitivity, spec=specificity, NPV=negative predictive value, PPV=positive predictive value. MFQ=mood and feelings questionnaire, FH=family history, CASE= Child and Adolescent Survey of Experiences **Definitions:** micro average is how well the model performs overall (weighted by group size), macro-average= model performance across all groups (not just largest)

A variety of supervised predictive modelling methods were used. These ranged from extensions of regression-based methods (e.g., LASSO) to computationally-advanced algorithms (e.g., Support Vector Machines). All development studies used internal cross-validation to assess model generalizability and attempt to reduce overfitting.

### Predictor/feature types, measurement, selection, and importance

The number of final predictors in models ranged from three to 152. Candidate predictor selection included literature based and statistically informed methods. Methods for final selection and determination of feature importance in the final model varied and were not always clear. Some studies used all predictors available in the dataset. Others used methods like recursive feature elimination to rank and remove less important features (Xiang et al., 2022). See supplementary information for complete predictor lists.

Models combined biological, psychological and sociodemographic predictors. Commonly used predictors were family psychiatric history (eight studies) (Dinga et al., 2018; Gorham et al., 2022; Kessler et al., 2016; Teutenberg et al., 2025; van Loo et al., 2014, 2014; Wardenaar et al., 2021; Xiang et al., 2022), features of incident/previous depression episode(s) (seven studies in subjects with baseline MDD) (Dinga et al., 2018; Gorham et al., 2022; Kessler et al., 2016; Teutenberg et al., 2025; van Loo et al., 2014; Wardenaar et al., 2021, 2014) and comorbid psychiatric conditions (six studies) (Dinga et al., 2018; Kessler et al., 2016; Schultebraucks et al., 2021; Teutenberg et al., 2025; van Loo et al., 2014; Wardenaar et al., 2021, 2014). Features from incident/previous depression episodes included age-of-onset and symptom profile. Other predictors included adverse childhood experiences (ACE) (five studies) (Dinga et al., 2018; Gorham et al., 2022; Teutenberg et al., 2025; Wardenaar et al., 2021; Xiang et al., 2022), sleep problems (five studies) (Kessler et al., 2016; van Loo et al., 2014; Wardenaar et al., 2021, 2014; Xiang et al., 2022) and educational factors (e.g., attainment) (four studies) (Dinga et al., 2018; Schultebraucks et al., 2021; Teutenberg et al., 2025; Wardenaar et al., 2021; Xiang et al., 2022) (Figure 2).

**Figure 2.**
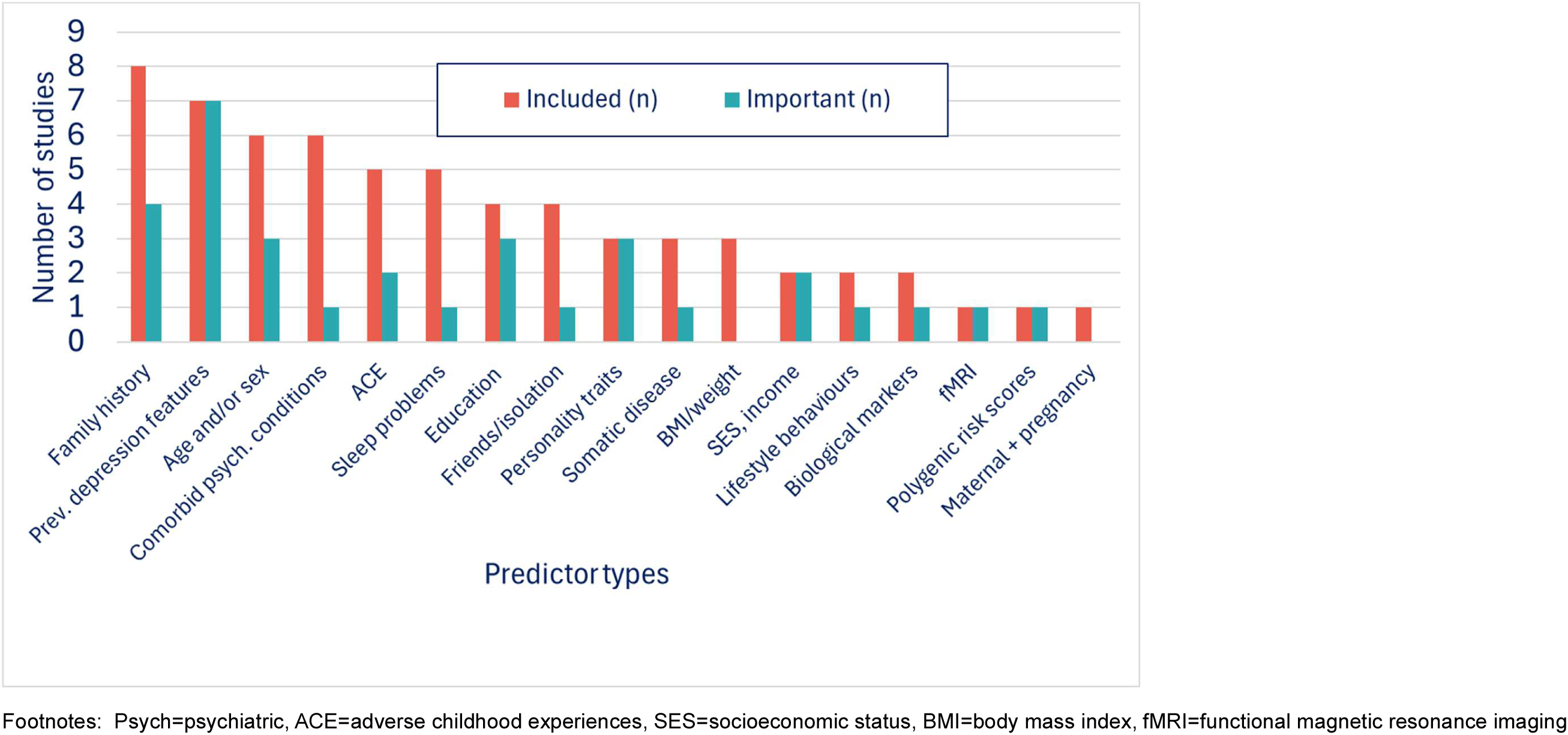
Predictor types, their frequency of use in prediction models, and feature importance across 9 studies reviewed.

Most studies measured predictors using questionnaire data or health care records. One study used polygenic scores (PGS) for phenotypic traits of common predictor types such as educational attainment (Schultebraucks et al., 2021). One study used fMRI data and maternal/pregnancy factors (Xiang et al., 2022).

Measurement of predictors varied between studies. For example, ACEs were measured by different questionnaires including the Child Trauma Questionnaire (CTQ) (Teutenberg et al., 2025) and the Child and Adolescent Survey of Experiences (CASE) (Gorham et al., 2022). The CTQ measures five core trauma types (emotional, physical and sexual abuse, emotional and physical neglect) (Bernstein et al., 1994). CASE includes additional items e.g., parental death (Allen and Rapee, 2012). Measurement of family psychiatric history also varied across studies. For example, some included only parental history, others included first- and second-degree relatives.

Predictors that were commonly used across studies did not always rank highly for predictive importance (Figure 2). For example, family psychiatric history, comorbid psychiatric conditions and ACEs were common predictors, but were reported to be important predictors in ≤50% of the models they featured in (Gorham et al., 2022; Teutenberg et al., 2025; Wardenaar et al., 2021). Conversely, personality traits (extroversion and neuroticism), and socioeconomic factors were important predictors in all models they were used (three and two studies respectively). Factors relating to incident/past depression were both commonly used (seven studies) and important predictors in all seven studies.

Only personality related factors appeared to consistently predict a particular trajectory type across studies. In the studies where personality types were used as predictors, extroversion was a consistent predictor of recovery and resilience trajectories (Dinga et al., 2018; Schultebraucks et al., 2021; Teutenberg et al., 2025; Wardenaar et al., 2021), while neuroticism predicted multiple/different trajectories (e.g., chronic-high, resilience and recovery) (Schultebraucks et al., 2021; Wardenaar et al., 2021). Extroversion and neuroticism remained important predictors in models with a large number of final predictors (between 21-152) (Dinga et al., 2018; Schultebraucks et al., 2021; Teutenberg et al., 2025; Wardenaar et al., 2021).

### Model performance

The best performing model predicted depression trajectories over a two year horizon in healthy adults aged 50+ following a single major life stressor, using data from the Health and Retirement study (Schultebraucks et al., 2021). Discriminative ability was high for multi-class discrimination of depression trajectories and individual trajectory group discrimination (all but one AUC>0.85). Model performance was highest for predicting chronic trajectory group membership (vs. all others) (AUC=0.93) and lowest for the resilience trajectory group (AUC=0.75) (Table 3). The micro-average AUC for multi-class discrimination was 0.88 (95%CI:0.87-0.89) with the macro-average dropping only slightly to 0.86 (95%CI:0.85-0.87). The model had good specificity (82%) and precision (79%) but lower sensitivity (recall=60%, 40% of true positives are missed). In this model, PGS were used as predictors and the most important predictor of chronic depression was PGS for high body fat distribution (negatively associated) followed by PGS for low wellbeing score, higher baseline depressive symptoms, low educational attainment and low neuroticism which were all positively associated with the chronic trajectory (Schultebraucks et al., 2021).

One other model reported performance for multi-class discrimination of depression trajectories, this was in a healthy population sample of children aged 9-10 years from the ABCD cohort (Xiang et al., 2022). The micro-average AUC was 0.90, indicating excellent discrimination. However, the macro-average AUC dropped to 0.77 suggesting class imbalance. AUCs for individual trajectory group predictions are not reported. This model had good specificity (82%) but poor sensitivity (recall=45%) and precision (45%). Sleep disturbance, family financial adversities in the past year and parent/adult self-report psychopathology scores were important predictors (de Vries et al., 2020). Among the top 10 predictors was fMRI data of correlations between attention networks and other brain regions.

Two studies (Dinga et al., 2018; Teutenberg et al., 2025) compared the performance of models incorporating different predictor groups. A model incorporating 18 biological measures performed worse than a model incorporating five personality traits and a model combining 55 “clinical” predictors (predictors such as family psychiatric history and ACE). Performance only slightly differed across models. AUCs were <70 for all models (range 0.56-0.69) (supplement, sTable 7), including an overall model combining all biological, personality and clinical markers. Biological measures included both health-related anthropometric information (e.g., BMI) and blood inflammatory markers (e.g., interleukin-6 [IL-6] and C-reactive protein [CRP]).

Teutenberg *et al*. (2025) (Teutenberg et al., 2025) compared three models which grouped predictors based on prior evidence of their strength as prediction factors for depression. Predictors with “strong evidence” included ACEs. “Good evidence” predictors included neuroticism. “Exploratory” predictors included family psychiatric history and extraversion. However, the results showed that “strong evidence” predictors do not necessarily perform well; the “strong evidence” model performed the worst for predicting all trajectories (Table 3, supplementary table 7).

Two models report performance as mean-squared errors (MSE) and do not report AUCs (Gorham et al., 2022; Wardenaar et al., 2021). Therefore, model performance cannot be directly compared. One paper also gives correlations of predicted-to-observed (o/e) scores (similar to the brier score measure of calibration). The smallest MSE (smaller=less predictive error) was observed in the full-recovery depression trajectory and increasing severity of anxiety groups. However the best o/e score was observed in the partial-recovery groups (depression and anxiety) (Wardenaar et al., 2021).

Overall, there were no clear patterns across studies suggesting that models were consistently able to discriminate a specific trajectory group (e.g., chronic) better than any other group.

### Risk of bias assessment

Overall RoB was high in all studies (Table 4, supplementary Table s5: full assessment). Due to the longitudinal design of most studies, a common area of low concern was timing of predictor measurement (at baseline, temporally preceding outcome measurement). However, all studies had high RoB in analysis and outcome domains. It was often unclear if predictor data was collected without knowledge of the outcome (“blinded”), due to self-reported or retrospective outcome data collection.

**Table 4.** Risk of bias in studies reviewed.

| Author, Year | Risk of Bias |  |  |  | Applicability |  |  | Overall |  |
| --- | --- | --- | --- | --- | --- | --- | --- | --- | --- |
|  | 1. Participants | 2. Predictors | 3. Outcome | 4. Analysis | 1. Participants | 2. Predictors | 3. Outcome | Risk of Bias | Applicability |
| Teutenberg, 2025 | - | + | - | - | - | + | + | - | - |
| Gorham, 2022 | - | + | - | - | + | + | - | - | - |
| Xiang, 2022 | + | + | - | - | + | - | + | - | - |
| Schultebrasucks, 2021 | - | + | - | - | - | - | + | - | - |
| Wardenaar, 2021 | - | + | - | - | - | + | + | - | - |
| Dinga, 2018 | - | + | - | - | - | + | + | - | - |
| Kessler, 2016 | + | - | - | - | + | + | - | - | - |
| Van Loo, 2014 | - | - | - | - | + | + | - | - | - |
| Wardenaar, 2014 | - | - | - | - | + | + | - | - | - |

Footnote: + (green) = low risk of bias concern, - (red) = high risk of bias concern

A universal source of bias was potential model overfitting due to small samples, combined with many predictive factors and computationally intensive machine learning methods, which typically require large samples (Riley et al., 2025). Only one included study discussed adequacy of the overall sample (Gorham et al., 2022). We calculated events-per-variable (EPV) based on the information provided in the papers (supplementary sTables 3-5). EPV ranged from 0.64 to 4.34 in all multinomial and binomial models. Model overfitting could be a reason for apparent superior model performance in studies using class-based trajectory modelling techniques (Dinga et al., 2018; Schultebraucks et al., 2021; Teutenberg et al., 2025; Wardenaar et al., 2021; Xiang et al., 2022), especially in chronic/persistent trajectory groups because these had the smallest samples. Small samples contribute to other sources of bias e.g., lack of generalizability. All studies performed internal cross-validation to assess for overfitting and model optimism, but it was not always clear whether that led to subsequent adjustment. No studies reported both discrimination and calibration measures.

Generally, applicability ratings were better than RoB. However, at least one applicability domain was rated as high concern for each study, leading to a high overall rating. In four studies (Gorham et al., 2022; Kessler et al., 2016; van Loo et al., 2014; Wardenaar et al., 2014) this was because unconventional trajectory modelling methods were used. In four studies, concern was related to applicability of study samples to our specified general population criteria (Dinga et al., 2018; Schultebraucks et al., 2021; Teutenberg et al., 2025; Wardenaar et al., 2021). In three of these, subjects had baseline MDD and were from cohorts sampling the general population, primary care, and secondary care (Dinga et al., 2018; Teutenberg et al., 2025; Wardenaar et al., 2014). Because the subjects had MDD at baseline, a higher proportion may be from secondary care than the general population.

## 4. Discussion

To our knowledge, this is the first systematic review of prediction models for forecasting longitudinal trajectories of depression and anxiety. Nine population-based studies were included, mostly predicting depression trajectories in adults with baseline depression. We found only one study predicting anxiety trajectories, and only two studies in child/adolescent populations. Only one model had an associated external validation study, but this used different predictors to the development models. Multinominal models performed better than those predicting individual trajectories. Models generally had good specificity but poor sensitivity.

Models combined biological, psychological and sociodemographic predictors. Family and personal psychiatric history were the most common predictors. Features of past depressive episodes were important predictors and extroversion was a consistent predictor of recovery/resilience trajectories. However, there were no other clear patterns suggesting that individual predictors or models consistently discriminated a particular trajectory group from others.

While our findings suggest prediction of future trajectories appears to be feasible, external validation in large independent samples and assessment of clinical utility are needed. Exploring novel predictors may improve future model performance.

### Findings in context of existing literature

Previous systematic reviews have identified prediction models for anxiety and depression but most have focused on binary outcomes, such as diagnosis or relapse by a single timepoint, in depressed adults (Meehan et al., 2022; Moriarty et al., 2022; Senior et al., 2021; Todd et al., 2025). Other reviews have identified studies modelling depression and anxiety trajectories in diverse population groups (Musliner et al., 2016; Shore et al., 2018). However, we lack multivariable prediction models to forecast these trajectories.

Our review builds on existing research by summarizing the literature on prediction models that predict longitudinal trajectories of depression/anxiety. We identify a need for models that can forecast trajectories, especially in young people and general populations. Models for anxiety are lacking which is notable in young people given that 40% of cases manifest before age 14 (Solmi et al., 2022) and early onset can predict later depression (Davies et al., 2016). Models for predicting trajectories could help identify individuals at risk of worsening or persistent symptoms early, before symptoms escalate.

Our findings suggest it is possible to develop a model with reasonable predictive accuracy using different predictor types. However, models need to be improved before they can support clinical decision making. Like in previous reviews (Moriarty et al., 2022; Senior et al., 2021), no included models had undergone external validation and all studies were assessed as having high risk of bias. Therefore, model performance in novel samples may be overestimated and lack generalizability.

For models that reported discrimination, AUCs ranged from 60–75%. This range aligns with models identified by previous reviews predicting non-trajectory based depression/anxiety outcomes (Meehan et al., 2022; A. S. Moriarty et al., 2022; Senior et al., 2021). An AUC of 70% is generally considered to be the lower acceptable performance limit for prediction models (White et al., 2023). Clinically acceptable thresholds for predicting depression/anxiety need to be agreed given potential harms associated with incorrect prediction of depression and anxiety.

Included models had high specificity but low sensitivity which is consistent with previous research (Moriarty et al., 2022; Senior et al., 2021; Todd et al., 2025). This means models more reliably identify people who *won’t* develop persistent symptoms, rather than those who will. Good sensitivity may have advantages, such as avoidance of unnecessary intervention or stigma. Acceptable clinical thresholds need to be agreed for sensitivity and specificity before models can be implemented.

One model had excellent performance. However, this could be due to overfitting to the data and specific study population (healthy older adults after a major life stressor). The model needs to be validated in external datasets to better understand performance. This model used genetic data which is becoming more common in prediction research. Novel machine learning techniques have computational power to handle a large number of complex predictors (like genome-wide data). However, we found that models with more predictors did not always have superior performance and some excluded useful, accessible predictors like sociodemographic features. Previous studies suggest that routinely available predictors (e.g., family history and sociodemographic factors) may have better predictive value than genetic data (Allesøe et al., 2023).

Common predictors identified in this review align with previous evidence for individual predictive factors of depression course (Buckman et al., 2018; Tagliaferri et al., 2025). However, common predictors based on strong prior evidence were not always the most important contributors to model performance. This could be because predictive importance depends on factors such as sample size and the other predictors included in a model; a predictor may appear relatively less important in models with many variables vs a model with fewer variables. Despite this, we found that neuroticism and extraversion were consistently important even in models with ~80-150 predictors (Dinga et al., 2018; Schultebraucks et al., 2021; Teutenberg et al., 2025; Wardenaar et al., 2021). One possible explanation for this is that extroversion/neuroticism capture traits overlapping with depression and anxiety (e.g., worry, social withdrawal).

### Implications for future research, policy and practice

Our findings do not suggest that adding difficult-to-measure predictors provides additional benefit. In future studies, balancing model complexity and clinical utility will be key as specialist predictors, such as neuroimaging data, are costly and not routinely available. Adding a large number of predictors without considering sample size can compromise model quality and utility. Simple models have shown promise. For example, the “IDEA-RS” model combines 11 accessible sociodemographic factors and has performed well in development and validation studies (Kieling et al., 2021; Piccin et al., 2025; Rocha et al., 2021) for predicting any episode of depression by late adolescence.

Using routinely collected data and electronic records could automate risk scoring which might ultimately mean models could be implemented without increasing burden on health systems. Wearable and smart devices are ubiquitous (especially in young people) and can also provide easy-to-measure information for prediction. Future research should consider using such data (Fried et al., 2023).

While features of previous depression episodes were informative predictors, these are less relevant for predicting onset and trajectories in young people or healthy general populations. In these groups, subclinical symptoms, peer relationships and school engagement are examples of potentially more useful predictors, and are likely more readily available in community-based settings. Population-based birth cohort studies follow children from early life and collect rich health, social, and educational data, and are valuable resources for developing models to predict trajectories.

### Strengths and limitations

Strengths of our work include a rigorous, transparent review process. We followed current guidelines for systematic reviews of prediction models, used sensitive search terms, performed double screening, and dual data extraction. Nevertheless, the small number of heterogeneous studies limited conclusions and meant meta-analysis was not possible (Damen et al., 2023).

We rated all studies as “high concern” for applicability to the current review, which is a limitation. This was due to variation in trajectory modelling approaches; our definition of trajectories was broad and we included four studies that used unconventional methods to model trajectories (Gorham et al., 2022; Kessler et al., 2016; van Loo et al., 2014; Wardenaar et al., 2014). Findings for these four studies were difficult to compare with others. Synthesizing and comparing results across studies was challenging due to heterogeneity in other areas. For example: reporting of model performance was inconsistent (studies used different metrics, not all report discrimination and calibration measures), different models used different predictor sets (number, type and measures used) and different studies used different feature importance ranking methods. To our knowledge no method exists to create universal feature importance scores that are directly comparable across models.

The included studies did not always adhere to reporting guidelines for prediction models (TRIPOD-AI) (Collins et al., 2024). Lack of rationale/justification for sample size was an important gap in all studies and only one discussed this limitation (Gorham et al., 2022). Using inadequate sample sizes to train and validate models risk of overfitting, especially for those predicting chronic/persistent high symptom trajectory groups which had small samples. This is a common limitation of AI and machine learning models (de Jong et al., 2021; Riley et al., 2025), and means models may not generalize to the target population (especially for under-represented groups). Ultimately, this may lead to harmful and incorrect clinical decision making (Riley et al., 2025), and clinical utility of the models was not assessed.

## 5. Conclusions

This review highlights a need for robust prediction models to forecast future risk of persistent or worsening anxiety and depression, especially in child/adolescent populations where there is opportunity for early intervention. Studies in adults with major depression indicate that family and personal psychiatric history are important for predicting future depression trajectories. However, to what extent these are applicable to young people with emerging symptoms, and in predicting anxiety trajectories, requires further investigation.

## Contributions

SJF: Conceptualization, formal analysis (searches and data extraction), methodology, visualisation, writing – original draft preparation and editing. NL: methodology (protocol review), analysis (double screening abstracts), HF: analysis (double screening abstracts and data extraction checks), reviewing and editing. GMK: Conceptualization, methodology, supervision, writing – reviewing and editing. HJJ: methodology (protocol review), supervision, analysis (double screening abstracts and data extraction checks), writing – reviewing and editing. LP: methodology (protocol review), reviewing and editing. SG: conceptualization, writing – reviewing and editing.

## 7. Acknowledgements and funding

This publication is the work of the authors and Sophie Fairweather and Hannah Jones will serve as guarantors for the contents of this paper. This work was supported by the National Institute for Health and Care Research (NIHR) Bristol Biomedical Research Centre (SJF, HJJ, and GMK; grant no: NIHR 203315). SJF is supported by a NIHR Bristol Biomedical Research Centre PhD studentship. The views expressed are those of the authors and not necessarily those of the NIHR or the Department of Health and Social Care. GMK acknowledges funding support from the UK Medical Research Council (MRC), grant no: MC_UU_00032/6, which forms part of the Integrative Epidemiology Unit at the University of Bristol. GMK acknowledges additional funding from the Wellcome Trust (201486/Z/16/Z and 201486/B/16/Z), the MRC (MR/W014416/1; MR/S037675/1; and MR/Z50354X/1).

SG is supported by the NIHR Applied Research Collaboration [ARC] for Yorkshire and Humberside [reference number NIHR200166].

## 8. Data availability statement

All data from generated or analysed during this study are included in the cited, published articles for the nine included studies [and their] supplementary information files(Dinga et al., 2018; Gorham et al., 2022; Kessler et al., 2016; Schultebraucks et al., 2021; Teutenberg et al., 2025; van Loo et al., 2014; Wardenaar et al., 2021, 2014; Xiang et al., 2022).

