## Supplementary material for "Prediction models for longitudinal trajectories of depression and anxiety: a systematic review"

### Contents

Supplementary methods ... 3
Supplementary Table 1. PRISMA Checklist ... 3
Supplementary Table 2. PRISMA Checklist for abstracts ... 7
Full eligibility criteria ... 8
Full search strategy ... 10
Reasons for full text exclusions (PRISMA item 16b) ... 22
Additional data extraction methods ... 26
Additional risk of bias assessment details ... 26
PROBAST guidance from Moons et al., 2025 ... 26
EPV calculations ... 27
Supplementary Table 3. EPV calculations for multinomial models ... 28
Supplementary table 4. EPV calculations for binomial one-vs-rest models ... 28
Supplementary table 5. EPV calculations for continuous outcomes ... 29
Minor protocol deviations ... 30
Supplementary data extraction ... 30
Search results and reasons for exclusions ... 30
Interpretation of methods: Kessler 2016, Van Loo and Wardenaar 2014 ... 31
Detailed data extraction tables ... 31
Xiang 2022 ... 31
Wardenaar 2021 ... 33
Dinga 2018 ... 35
*Kessler 2016* ... 38
Van Loo 2014 ... 40
Wardenaar 2014 ... 42
Gorham 2022 ... 44
Schultebraucks 2021 ... 47
Teutenberg 2025 ... 49
Supplementary table 6. Number of unique models in each study ... 52
Supplementary table 7. Performance statistics of other models ... 53
Detailed risk of bias assessment for best performing models ... 55
Xiang 2022 ... 55
Wardenaar 2021 ... 57
Dinga 2018 ... 59
Kessler 2016 ... 61

### Supplementary methods

#### Supplementary Table 1. PRISMA Checklist

| Section and Topic | Item # | Checklist item | Location where item is reported |
|---|---|---|---|
| **TITLE** | | | |
| Title | 1 | Identify the report as a systematic review. | Main text - Title |
| **ABSTRACT** | | | |
| Abstract | 2 | See the PRISMA 2020 for Abstracts checklist. | See table below for PRISMA abstract checklist. |
| **INTRODUCTION** | | | |
| Rationale | 3 | Describe the rationale for the review in the context of existing knowledge. | Main text - introduction – paragraphs 3 and 4 in particular |
| Objectives | 4 | Provide an explicit statement of the objective(s) or question(s) the review addresses. | Main text - end of introduction section |
| **METHODS** | | | |
| Eligibility criteria | 5 | Specify the inclusion and exclusion criteria for the review and how studies were grouped for the syntheses. | Main text - materials and methods section under heading “eligibility criteria, including table 1, and supplement for full list of eligibility |
| Information sources | 6 | Specify all databases, registers, websites, organisations, reference lists and other sources searched or consulted to identify studies. Specify the date when each source was last searched or consulted. | Main text - materials and methods section under heading “search strategy and databases” |
| Search strategy | 7 | Present the full search strategies for all databases, registers and websites, including any filters and limits used. | Supplement. |
| Selection process | 8 | Specify the methods used to decide whether a study met the inclusion criteria of the review, including how many reviewers screened each record and each report retrieved, whether they worked independently, and if applicable, details of automation tools used in the process. | Main text - materials and methods section under heading “study selection” |
| Data collection process | 9 | Specify the methods used to collect data from reports, including how many reviewers collected data from each report, whether they worked independently, any processes for obtaining or confirming data from study investigators, and if applicable, details of automation tools used in the process. | Main text - materials and methods section under heading “data extraction |

| Section and Topic | Item # | Checklist item | Location where item is reported |
|---|---|---|---|
| | | | and risk of bias…”. Further detail in supplement. |
| Data items | 10a | List and define all outcomes for which data were sought. Specify whether all results that were compatible with each outcome domain in each study were sought (e.g. for all measures, time points, analyses), and if not, the methods used to decide which results to collect. | Main text - materials and methods section (inc. Table 1), supplement. Best performing model reported in main text, additional models in supplement. |
| | 10b | List and define all other variables for which data were sought (e.g. participant and intervention characteristics, funding sources). Describe any assumptions made about any missing or unclear information. | Full list in supplement. CHARMS tool referenced in main text. |
| Study risk of bias assessment | 11 | Specify the methods used to assess risk of bias in the included studies, including details of the tool(s) used, how many reviewers assessed each study and whether they worked independently, and if applicable, details of automation tools used in the process. | Main text - materials and methods section under heading “data extraction and risk of bias…”. Further detail in supplement. PROBAST-AI tool used. |
| Effect measures | 12 | Specify for each outcome the effect measure(s) (e.g. risk ratio, mean difference) used in the synthesis or presentation of results. | Main text - materials and methods (inc. Table 1), supplement. We report performance measures of the included prediction models (any measure given). |
| Synthesis methods | 13a | Describe the processes used to decide which studies were eligible for each synthesis (e.g. tabulating the study intervention characteristics and comparing against the planned groups for each synthesis (item #5)). | n/a – all 9 studies in one table. |
| | 13b | Describe any methods required to prepare the data for presentation or synthesis, such as handling of missing summary statistics, or data conversions. | See supplementary section relating to EPV calculations. |
| | 13c | Describe any methods used to tabulate or visually display results of individual studies and syntheses. | Described in main text, materials and methods section under heading “data extraction and risk of bias…”. |

| Section and Topic | Item # | Checklist item | Location where item is reported |
|---|---|---|---|
| | 13d | Describe any methods used to synthesize results and provide a rationale for the choice(s). If meta-analysis was performed, describe the model(s), method(s) to identify the presence and extent of statistical heterogeneity, and software package(s) used. | n/a – we provide a rationale for why meta-analysis was not appropriate in the main text. |
| | 13e | Describe any methods used to explore possible causes of heterogeneity among study results (e.g. subgroup analysis, meta-regression). | n/a |
| | 13f | Describe any sensitivity analyses conducted to assess robustness of the synthesized results. | n/a |
| Reporting bias assessment | 14 | Describe any methods used to assess risk of bias due to missing results in a synthesis (arising from reporting biases). | n/a |
| Certainty assessment | 15 | Describe any methods used to assess certainty (or confidence) in the body of evidence for an outcome. | n/a |
| **RESULTS** | | | |
| Study selection | 16a | Describe the results of the search and selection process, from the number of records identified in the search to the number of studies included in the review, ideally using a flow diagram. | Main text – Figure 1. |
| | 16b | Cite studies that might appear to meet the inclusion criteria, but which were excluded, and explain why they were excluded. | Supplementary materials. |
| Study characteristics | 17 | Cite each included study and present its characteristics. | Main text – Table 2 and accompanying narrative. |
| Risk of bias in studies | 18 | Present assessments of risk of bias for each included study. | Supplementary materials. |
| Results of individual studies | 19 | For all outcomes, present, for each study: (a) summary statistics for each group (where appropriate) and (b) an effect estimate and its precision (e.g. confidence/credible interval), ideally using structured tables or plots. | Main text tables 3 and 4. Supplementary table 7. Confidence intervals not reported in some studies. |
| Results of syntheses | 20a | For each synthesis, briefly summarise the characteristics and risk of bias among contributing studies. | n/a – single narrative |
| | 20b | Present results of all statistical syntheses conducted. If meta-analysis was done, present for each the summary estimate and its precision (e.g. confidence/credible interval) and measures of statistical heterogeneity. If comparing groups, describe the direction of the effect. | n/a |
| | 20c | Present results of all investigations of possible causes of heterogeneity among study results. | n/a |
| | 20d | Present results of all sensitivity analyses conducted to assess the robustness of the synthesized results. | n/a |
| Reporting biases | 21 | Present assessments of risk of bias due to missing results (arising from reporting biases) for each synthesis assessed. | n/a |

| Section and Topic | Item # | Checklist item | Location where item is reported |
|---|---|---|---|
| Certainty of evidence | 22 | Present assessments of certainty (or confidence) in the body of evidence for each outcome assessed. | n/a |
| **DISCUSSION** | | | |
| Discussion | 23a | Provide a general interpretation of the results in the context of other evidence. | Main text – discussion – under heading “findings in context of existing literature” |
| | 23b | Discuss any limitations of the evidence included in the review. | Main text – discussion – under heading “strengths and limitations” |
| | 23c | Discuss any limitations of the review processes used. | Main text – discussion – under heading “strengths and limitations” |
| | 23d | Discuss implications of the results for practice, policy, and future research. | Main text – discussion – under heading “implications for future research and clinical practice” |
| **OTHER INFORMATION** | | | |
| Registration and protocol | 24a | Provide registration information for the review, including register name and registration number, or state that the review was not registered. | Main text – first paragraph od materials and methods. PROSPERO ID also given at end of abstract. |
| | 24b | Indicate where the review protocol can be accessed, or state that a protocol was not prepared. | Main text – first paragraph od materials and methods. PROSPERO ID also given at end of abstract. |
| | 24c | Describe and explain any amendments to information provided at registration or in the protocol. | Supplementary materials describes deviations from the protocol. We reference this in the main text. |
| Support | 25 | Describe sources of financial or non-financial support for the review, and the role of the funders or sponsors in the review. | Section 7 of main text – acknowledgements and funding. |

| Section and Topic | Item # | Checklist item | Location where item is reported |
|---|---|---|---|
| Competing interests | 26 | Declare any competing interests of review authors. | None to declare (stated under section 6 of main text). |
| Availability of data, code and other materials | 27 | Report which of the following are publicly available and where they can be found: template data collection forms; data extracted from included studies; data used for all analyses; analytic code; any other materials used in the review. | Supplement is referenced throughout main text. CHARMS and PROBAST templates referenced. Full data extraction provided in supplement. No analytic code used. |

*From:* Page MJ, McKenzie JE, Bossuyt PM, Boutron I, Hoffmann TC, Mulrow CD, et al. The PRISMA 2020 statement: an updated guideline for reporting systematic reviews. BMJ 2021;372:n71. doi: 10.1136/bmj.n71. This work is licensed under CC BY 4.0. To view a copy of this license, visit https://creativecommons.org/licenses/by/4.0/

### Supplementary Table 2. PRISMA Checklist for abstracts

| Section and Topic | Item # | Checklist item | Reported (Yes/No) |
|---|---|---|---|
| **TITLE** | | | |
| Title | 1 | Identify the report as a systematic review. | YES |
| **BACKGROUND** | | | |
| Objectives | 2 | Provide an explicit statement of the main objective(s) or question(s) the review addresses. | YES |
| **METHODS** | | | |
| Eligibility criteria | 3 | Specify the inclusion and exclusion criteria for the review. | YES |
| Information sources | 4 | Specify the information sources (e.g. databases, registers) used to identify studies and the date when each was last searched. | YES |
| Risk of bias | 5 | Specify the methods used to assess risk of bias in the included studies. | YES |
| Synthesis of results | 6 | Specify the methods used to present and synthesise results. | YES |
| **RESULTS** | | | |
| Included studies | 7 | Give the total number of included studies and participants and summarise relevant characteristics of studies. | YES |
| Synthesis of results | 8 | Present results for main outcomes, preferably indicating the number of included studies and participants for each. | YES |

| Section and Topic | Item # | Checklist item | Reported (Yes/No) |
|---|---|---|---|
| | | If meta-analysis was done, report the summary estimate and confidence/credible interval. If comparing groups, indicate the direction of the effect (i.e. which group is favoured). | |
| **DISCUSSION** | | | |
| Limitations of evidence | 9 | Provide a brief summary of the limitations of the evidence included in the review (e.g. study risk of bias, inconsistency and imprecision). | YES |
| Interpretation | 10 | Provide a general interpretation of the results and important implications. | YES |
| **OTHER** | | | |
| Funding | 11 | Specify the primary source of funding for the review. | YES |
| Registration | 12 | Provide the register name and registration number. | YES |

*From:* Page MJ, McKenzie JE, Bossuyt PM, Boutron I, Hoffmann TC, Mulrow CD, et al. The PRISMA 2020 statement: an updated guideline for reporting systematic reviews. BMJ 2021;372:n71. doi: 10.1136/bmj.n71. This work is licensed under CC BY 4.0. To view a copy of this license, visit https://creativecommons.org/licenses/by/4.0/

### Full eligibility criteria

#### Population

**Inclusion:**

- Longitudinal patterns (trajectories) of depression and anxiety from general population-based samples including child, adolescent, young adult and adult populations.
- Trajectories starting at any age between 3 years and 65 years (inclusive).

**Exclusion:**

- Children under 3 years and adults over the age of 65 years.
- Specialist populations for example, child mental health clinic attendees, survivors of an earthquake, individuals with specific comorbidities such as cancer or HIV.

#### Intervention(s) or exposure(s)

*In the context of a systematic review of prediction models, the intervention is the index model. We use the term "Prediction model" to refer to any study aimed at developing, validating, and adjusting (e.g. extending) multivariable prediction/prognostic model(s) that include multiple prediction/prognostic factors combined, and are to be used for making predictions in individuals.*

**Inclusion:**

- Any prediction model predicting longitudinal patterns of depression and/or anxiety diagnosis/symptoms (including studies which use pre-existing longitudinal trajectories, in which the focus is prediction and not on trajectory modelling).
- Studies of model development and/or validation and/or extensions of existing models.
- All/any types of predictors/factors from any age or timepoint are eligible for inclusion.
- Models using any modelling method e.g., statistical methods, and machine learning methods. Supervised and unsupervised.

**Exclusion:**

- Single "risk-factor" or prognostic factor regression models which only report coefficients or effect estimates from the regression models (e.g., as odds or risk ratios) rather than prediction-based performance indicators (i.e., discrimination and calibration).
- Development studies with no internal validation step.

#### Comparator(s) or control(s)

**Inclusion:**

- Studies relating to a single prediction model and/or studies comparing multiple prediction models will be included (for example, a study may compare the performance of a simplified model with a more complex model including additional predictive factors).
- Studies comparing/validating the same model in different populations (e.g., different age groups) or settings are also eligible if they fit with all other inclusion criteria.

**Exclusion:**

- Studies solely comparing change in anxiety/depression trajectories before and after an intervention (i.e., treatment response trajectories).

#### Context

- Studies not based on general population-based samples, e.g., those conducted in specialist settings with specialist populations such as child mental health clinic attendees or individuals with specific comorbidities such as cancer patients.

#### Full search strategy

This search strategy was validated by confirming that it retrieved key papers (already known to lead author) which matched the search criteria. The strategy was developed with input from the Population Health Sciences subject librarian at the University of Bristol:

MEDLINE

1. Predict* model.mp.
2. prognos* model.mp.
3. prognos* score.mp.
4. risk predict*.mp.
5. risk score.mp.
6. risk calculat*.mp.
7. risk model.mp.
8. risk index.mp.
9. risk assessment.mp.
10. risk scor*.mp.
11. prognostic scor*.mp.
12. predict* score.mp.
13. prediction scor*.mp.
14. predictive scor*.mp.
15. predict* index.mp.

16. prognos* index.mp.
17. prognostic model*.mp.
18. predictive model*.mp.
19. predict* instrument.mp.
20. prognos* instrument.mp.
21. predict* algorithm.mp.
22. prognos* algorithm.mp.
23. risk tool$.mp.
24. (risk adj3 tool).mp.
25. trajector*.mp.
26. developmental pattern*.mp.
27. longitudinal pattern*.mp.
28. longitudinal chang*.mp.
29. growth curve model*.mp.
30. latent class analysis/
31. temporal pattern*.mp.
32. temporal chang*.mp.
33. growth mixture model*.mp.
34. Depression/
35. depress*.mp.

36. depressive disorder/ or depressive disorder, major/
37. persistent depress*.mp.
38. Anxiety Disorders/
39. Anxiety/
40. generali?ed anxiety.mp.
41. atypical depression.mp.
42. ("Anxiety, Separation" or "Neurotic Disorders" or "Obsessive-Compulsive Disorder" or "Panic Disorder" or "Phobic Disorders").mp.
43. "Depression (Emotion)".mp.
44. ("Generalized Anxiety Disorder" or "Separation Anxiety Disorder").mp.
45. (predict* adj3 tool).mp.
46. (predict* or forecast*).mp.
47. exp Anxiety Disorders/ep
48. exp Anxiety Disorders/di
49. exp Depression/ep
50. exp Depression/di
51. 1 or 2 or 3 or 4 or 5 or 6 or 7 or 8 or 9 or 10 or 11 or 12 or 13 or 14 or 15 or 16 or 17 or 18 or 19 or 20 or 21 or 22 or 23 or 24 or 45 or 46
52. 25 or 26 or 27 or 28 or 29 or 30 or 31 or 32 or 33
53. 34 or 35 or 36 or 37 or 38 or 39 or 40 or 41 or 42 or 43 or 44 or 47 or 48 or 49 or 50
54. ((trajector* or developmental pattern* or longitudinal pattern* or longitudinal chang* or growth curve model*).mp. or latent class analysis/ or temporal pattern*.mp. or temporal chang*.mp. or growth mixture model*.mp.) adj6 (Depression/ or depress*.mp. or (depressive disorder/ or depressive disorder, major/) or persistent depress*.mp. or Anxiety Disorders/ or Anxiety/ or generali?ed anxiety.mp. or atypical depression.mp. or ("Anxiety, Separation" or "Neurotic Disorders" or "Obsessive-Compulsive Disorder" or "Panic Disorder" or

"Phobic Disorders").mp. or "Depression (Emotion)".mp. or ("Generalized Anxiety Disorder" or "Separation Anxiety Disorder").mp. or exp Anxiety Disorders/ep or exp Anxiety Disorders/di or exp Depression/ep or exp Depression/di)

55. (Predict* model or prognos* model or prognos* score or risk predict* or risk score or risk calculat* or risk model or risk index or risk assessment or risk scor* or prognostic scor* or predict* score or prediction scor* or predictive scor* or predict* index or prognos* index or prognostic model* or predictive model* or predict* instrument or prognos* instrument or predict* algorithm or prognos* algorithm or risk tool$ or (risk adj3 tool) or (predict* adj3 tool) or (predict* or forecast*)).mp. adj6 (((trajector* or developmental pattern* or longitudinal pattern* or longitudinal chang* or growth curve model*).mp. or latent class analysis/ or temporal pattern*.mp. or temporal chang*.mp. or growth mixture model*.mp.) adj6 (Depression/ or depress*.mp. or (depressive disorder/ or depressive disorder, major/) or persistent depress*.mp. or Anxiety Disorders/ or Anxiety/ or generali?ed anxiety.mp. or atypical depression.mp. or ("Anxiety, Separation" or "Neurotic Disorders" or "Obsessive-Compulsive Disorder" or "Panic Disorder" or "Phobic Disorders").mp. or "Depression (Emotion)".mp. or ("Generalized Anxiety Disorder" or "Separation Anxiety Disorder").mp. or exp Anxiety Disorders/ep or exp Anxiety Disorders/di or exp Depression/ep or exp Depression/di))
56. limit 55 to english language
57. remove duplicates from 56

Embase

1. Predict* model.mp.
2. prognos* model.mp.
3. prognos* score.mp.
4. risk predict*.mp.
5. risk score.mp.
6. risk calculat*.mp.
7. risk model.mp.
8. risk index.mp.

9. risk assessment.mp.
10. risk scor*.mp.
11. prognostic scor*.mp.
12. predict* score.mp.
13. prediction scor*.mp.
14. predictive scor*.mp.
15. predict* index.mp.
16. prognos* index.mp.
17. prognostic model*.mp.
18. predictive model*.mp.
19. predict* instrument.mp.
20. prognos* instrument.mp.
21. predict* algorithm.mp.
22. prognos* algorithm.mp.
23. risk tool$.mp.
24. (risk adj3 tool).mp.
25. trajector*.mp.
26. developmental pattern*.mp.
27. longitudinal pattern*.mp.
28. longitudinal chang*.mp.

29. growth curve model*.mp.
30. latent class analysis/
31. temporal pattern*.mp.
32. temporal chang*.mp.
33. growth mixture model*.mp.
34. Depression/
35. depress*.mp.
36. depressive disorder/ or depressive disorder, major/
37. persistent depress*.mp.
38. Anxiety Disorders/
39. Anxiety/
40. generali?ed anxiety.mp.
41. atypical depression.mp.
42. ("Anxiety, Separation" or "Neurotic Disorders" or "Obsessive-Compulsive Disorder" or "Panic Disorder" or "Phobic Disorders").mp.
43. "Depression (Emotion)".mp.
44. ("Generalized Anxiety Disorder" or "Separation Anxiety Disorder").mp.
45. exp prediction/
46. predict* analysis.mp.
47. Predict* model*.mp.
48. treatment resistant depression/

49. chronic depression/
50. atypical depression/
51. long term depression/
52. adolescent depression/
53. minor depression/
54. major depression/
55. recurrent brief depression/
56. persistent depress*.mp.
57. exp social anxiety/
58. exp "mixed anxiety and depression"/
59. affective disorder*.mp.
60. mood disorder/
61. (predict* adj3 tool).mp.
62. (predict* or forecast*).mp.
63. exp Anxiety Disorders/ep or exp Anxiety Disorders/di
64. exp Depression/ep or exp Depression/di
65. 1 or 2 or 3 or 4 or 5 or 6 or 7 or 8 or 9 or 10 or 11 or 12 or 13 or 14 or 15 or 16 or 17 or 18 or 19 or 20 or 21 or 22 or 23 or 24 or 45 or 46 or 47 or 61 or 62
66. 25 or 26 or 27 or 28 or 29 or 30 or 31 or 32 or 33
67. 34 or 35 or 36 or 37 or 38 or 39 or 40 or 41 or 42 or 43 or 44 or 48 or 49 or 50 or 51 or 52 or 53 or 54 or 55 or 56 or 57 or 58 or 59 or 60 or 63 or 64

68. ((trajector* or developmental pattern* or longitudinal pattern* or longitudinal chang* or growth curve model*).mp. or latent class analysis/ or temporal pattern*.mp. or temporal chang*.mp. or growth mixture model*.mp.) adj6 (Depression/ or depress*.mp. or (depressive disorder/ or depressive disorder, major/) or persistent depress*.mp. or Anxiety Disorders/ or Anxiety/ or generali?ed anxiety.mp. or atypical depression.mp. or ("Anxiety, Separation" or "Neurotic Disorders" or "Obsessive-Compulsive Disorder" or "Panic Disorder" or "Phobic Disorders").mp. or "Depression (Emotion)".mp. or ("Generalized Anxiety Disorder" or "Separation Anxiety Disorder").mp. or treatment resistant depression/ or chronic depression/ or atypical depression/ or long term depression/ or adolescent depression/ or minor depression/ or major depression/ or recurrent brief depression/ or persistent depress*.mp. or exp social anxiety/ or exp "mixed anxiety and depression"/ or affective disorder*.mp. or mood disorder/ or (exp Anxiety Disorders/ep or exp Anxiety Disorders/di) or (exp Depression/ep or exp Depression/di))
69. ((Predict* model or prognos* model or prognos* score or risk predict* or risk score or risk calculat* or risk model or risk index or risk assessment or risk scor* or prognostic scor* or predict* score or prediction scor* or predictive scor* or predict* index or prognos* index or prognostic model* or predictive model* or predict* instrument or prognos* instrument or predict* algorithm or prognos* algorithm or risk tool$ or (risk adj3 tool)).mp. or exp prediction/ or predict* analysis.mp. or Predict* model*.mp. or (predict* adj3 tool).mp. or (predict* or forecast*).mp.) adj6 (((trajector* or developmental pattern* or longitudinal pattern* or longitudinal chang* or growth curve model*).mp. or latent class analysis/ or temporal pattern*.mp. or temporal chang*.mp. or growth mixture model*.mp.) adj6 (Depression/ or depress*.mp. or (depressive disorder/ or depressive disorder, major/) or persistent depress*.mp. or Anxiety Disorders/ or Anxiety/ or generali?ed anxiety.mp. or atypical depression.mp. or ("Anxiety, Separation" or "Neurotic Disorders" or "Obsessive-Compulsive Disorder" or "Panic Disorder" or "Phobic Disorders").mp. or "Depression (Emotion)".mp. or ("Generalized Anxiety Disorder" or "Separation Anxiety Disorder").mp. or treatment resistant depression/ or chronic depression/ or atypical depression/ or long term depression/ or adolescent depression/ or minor depression/ or major depression/ or recurrent brief depression/ or persistent depress*.mp. or exp social anxiety/ or exp "mixed anxiety and depression"/ or affective disorder*.mp. or mood disorder/ or (exp Anxiety Disorders/ep or exp Anxiety Disorders/di) or (exp Depression/ep or exp Depression/di)))
70. limit 69 to english language
71. remove duplicates from 70

APA Psych INFO

1. Predict* model.mp.
2. prognos* model.mp.

3. prognos* score.mp.
4. risk predict*.mp.
5. risk score.mp.
6. risk calculat*.mp.
7. risk model.mp.
8. risk index.mp.
9. risk assessment.mp.
10. risk scor*.mp.
11. prognostic scor*.mp.
12. predict* score.mp.
13. prediction scor*.mp.
14. predictive scor*.mp.
15. predict* index.mp.
16. prognos* index.mp.
17. prognostic model*.mp.
18. predictive model*.mp.
19. predict* instrument.mp.
20. prognos* instrument.mp.
21. predict* algorithm.mp.
22. prognos* algorithm.mp.

23. risk tool$.mp.
24. (risk adj3 tool).mp.
25. trajector*.mp.
26. developmental pattern*.mp.
27. longitudinal pattern*.mp.
28. longitudinal chang*.mp.
29. growth curve model*.mp.
30. latent class analysis/
31. temporal pattern*.mp.
32. temporal chang*.mp.
33. growth mixture model*.mp.
34. Depression/
35. depress*.mp.
36. depressive disorder/ or depressive disorder, major/
37. persistent depress*.mp.
38. Anxiety Disorders/
39. Anxiety/
40. generali?ed anxiety.mp.
41. atypical depression.mp.
42. ("Anxiety, Separation" or "Neurotic Disorders" or "Obsessive-Compulsive Disorder" or "Panic Disorder" or "Phobic Disorders").mp.

43. "Depression (Emotion)".mp.
44. ("Generalized Anxiety Disorder" or "Separation Anxiety Disorder").mp.
45. exp predictive analysis/
46. developmental pattern*.mp.
47. exp Recurrent Depression/
48. exp "Depression (Emotion)"/
49. exp Treatment Resistant Depression/
50. exp Major Depression/
51. exp Atypical Depression/
52. exp Reactive Depression/
53. exp Anxiety Screening/
54. exp Separation Anxiety Disorder/
55. exp Social Anxiety/
56. exp Generalized Anxiety Disorder/
57. exp Anxiety Sensitivity/
58. exp Separation Anxiety/
59. exp affective disorders/
60. youth mental health/
61. (predict* adj3 tool).mp.
62. (predict* or forecast*).mp.

63. course trajectory.mp.
64. 1 or 2 or 3 or 4 or 5 or 6 or 7 or 8 or 9 or 10 or 11 or 12 or 13 or 14 or 15 or 16 or 17 or 18 or 19 or 20 or 21 or 22 or 23 or 24 or 45 or 61 or 62
65. 25 or 26 or 27 or 28 or 29 or 30 or 31 or 32 or 33 or 46 or 63
66. 34 or 35 or 36 or 37 or 38 or 39 or 40 or 41 or 42 or 43 or 44 or 45 or 46 or 47 or 48 or 49 or 50 or 51 or 52 or 53 or 54 or 55 or 56 or 57 or 58 or 59 or 60
67. ((trajector* or developmental pattern* or longitudinal pattern* or longitudinal chang* or growth curve model*).mp. or latent class analysis/ or temporal pattern*.mp. or temporal chang*.mp. or growth mixture model*.mp. or developmental pattern*.mp. or course trajectory.mp.) adj6 (Depression/ or depress*.mp. or (depressive disorder/ or depressive disorder, major/) or persistent depress*.mp. or Anxiety Disorders/ or Anxiety/ or generali?ed anxiety.mp. or atypical depression.mp. or ("Anxiety, Separation" or "Neurotic Disorders" or "Obsessive-Compulsive Disorder" or "Panic Disorder" or "Phobic Disorders").mp. or "Depression (Emotion)".mp. or ("Generalized Anxiety Disorder" or "Separation Anxiety Disorder").mp. or exp predictive analysis/ or developmental pattern*.mp. or exp Recurrent Depression/ or exp "Depression (Emotion)"/ or exp Treatment Resistant Depression/ or exp Major Depression/ or exp Atypical Depression/ or exp Reactive Depression/ or exp Anxiety Screening/ or exp Separation Anxiety Disorder/ or exp Social Anxiety/ or exp Generalized Anxiety Disorder/ or exp Anxiety Sensitivity/ or exp Separation Anxiety/ or exp affective disorders/ or youth mental health/)
68. ((Predict* model or prognos* model or prognos* score or risk predict* or risk score or risk calculat* or risk model or risk index or risk assessment or risk scor* or prognostic scor* or predict* score or prediction scor* or predictive scor* or predict* index or prognos* index or prognostic model* or predictive model* or predict* instrument or prognos* instrument or predict* algorithm or prognos* algorithm or risk tool$ or (risk adj3 tool)).mp. or exp predictive analysis/ or (predict* adj3 tool).mp. or (predict* or forecast*).mp.) adj6 (((trajector* or developmental pattern* or longitudinal pattern* or longitudinal chang* or growth curve model*).mp. or latent class analysis/ or temporal pattern*.mp. or temporal chang*.mp. or growth mixture model*.mp. or developmental pattern*.mp. or course trajectory.mp.) adj6 (Depression/ or depress*.mp. or (depressive disorder/ or depressive disorder, major/) or persistent depress*.mp. or Anxiety Disorders/ or Anxiety/ or generali?ed anxiety.mp. or atypical depression.mp. or ("Anxiety, Separation" or "Neurotic Disorders" or "Obsessive-Compulsive Disorder" or "Panic Disorder" or "Phobic Disorders").mp. or "Depression (Emotion)".mp. or ("Generalized Anxiety Disorder" or "Separation Anxiety Disorder").mp. or exp predictive analysis/ or developmental pattern*.mp. or exp Recurrent Depression/ or exp "Depression (Emotion)"/ or exp Treatment Resistant Depression/ or exp Major Depression/ or exp Atypical Depression/ or exp Reactive Depression/ or exp Anxiety Screening/ or exp Separation Anxiety Disorder/ or exp Social Anxiety/ or exp Generalized Anxiety Disorder/ or exp Anxiety Sensitivity/ or exp Separation Anxiety/ or exp affective disorders/ or youth mental health/))

69. limit 68 to english language
70. remove duplicates from 69

### Reasons for full text exclusions (PRISMA item 16b)

**PRISMA item 16b: "Cite studies that might appear to meet the inclusion criteria, but which were excluded, and explain why they were excluded."**

**Reports assessed for eligibility (database search results):**

Reports excluded: (n=109)

- Not prediction model (n=64)
- Outcome not trajectories (n=25)
- Ineligible population group (n=14)
- Review/other (n=6)

Emailed author for further info, no response: (n= 1)

**Below is a list of full texts from the database search that were assessed for eligibility and subsequently excluded (n=109), with rationale for exclusion:**

| Author, Year | Primary reason for exclusion | Additional notes |
|---|---|---|
| 1. Bosman 2020 [1] | Not true multivariable prediction model | Did not include an internal validation step |
| 2. Chen 2022 [2] | Ineligible population group and setting | Specialist setting; COVID specifically in Hubei (China), predictors context specific |
| 3. Gonzalez Colom 2024 [3] | Outcome not trajectories of dep/anx | |
| 4. Mallett 2022 [4] | Not true multivariable prediction model | |
| 5. Wang 2022 [5] | Not true multivariable prediction model | |
| 6. Norona-Zhou 2021 [6] | Not true multivariable prediction model | Individual risk factor |
| 7. Verhoeven 2020 [7] | Not true multivariable prediction model | Did not include an internal validation step |
| 8. Fokkema 2020 [8] | Outcome not trajectories of dep/anx | |
| 9. McGiffin 2019 [9] | Not true multivariable prediction model | Individual risk factor and specific population group (disability) |

| | | |
|---|---|---|
| 10. Chondros 2018 [10] | Outcome not trajectories of dep/anx | |
| 11. Finan 2017 [11] | Not true multivariable prediction model | Individual risk factors |
| 12. Whalen 2016 [12] | Not true multivariable prediction model | |
| 13. Elovainio 2014 [13] | Not true multivariable prediction model | Individual risk factors |
| 14. Cumsille 2015 [14] | Not true multivariable prediction model | |
| 15. Schmaal 2014 [15] | Not true multivariable prediction model | Did not include an internal validation step |
| 16. Hsu 2011 [16] | Not true multivariable prediction model | & in “older population” |
| 17. Best 2011 | Not true multivariable prediction model | Did not include an internal validation step |
| 18. Spinhoven 2011 [17] | Not true multivariable prediction model | Individual risk factor |
| 19. Garber 2010 [18] | Not true multivariable prediction model | Did not include an internal validation step |
| 20. Stoolmiller 2005 [19] | Not true multivariable prediction model | Did not include an internal validation step |
| 21. Garber 2002 [20] | Not true multivariable prediction model | Did not include an internal validation step |
| 22. Gonzalez-Colom 2023 [21] | Outcome not trajectories of dep/anx | |
| 23. Shu 2023 [22] | Not true multivariable prediction model | |
| 24. Meisenzhal 2024 [23] | Not true multivariable prediction model | |
| 25. Lin 2023 [24] | Ineligible population group | Likely that majority of sample over 65. |
| 26. Klawohn 2022 [25] | Not true multivariable prediction model | |
| 27. Whatnall 2022 [26] | Outcome not trajectories of dep/anx | |
| 28. Emden 2021 [27] | Not true multivariable prediction model | |
| 29. Frassle 2021 [28] | Ineligible population group | Not general population |
| 30. Maddison 2018 [29] | Outcome not trajectories of dep/anx | |
| 31. Hebebrand 2013 [30] | Not true multivariable prediction model | |
| 32. Fernandez-Castelao 2013 [31] | Not true multivariable prediction model | |
| 33. Penninx 2011 [32] | Not true multivariable prediction model | Did not include an internal validation step |
| 34. Panaite 2020 [33] | Not true multivariable prediction model | |
| 35. Fisher 2020 [34] | Outcome not trajectories of dep/anx | |
| 36. Tilton Weaver 2019 [35] | Not true multivariable prediction model | |
| 37. Timm 2017 [36] | Not true multivariable prediction model | Did not include an internal validation step |
| 38. Lamers 2016 | Not true multivariable prediction model | Did not include an internal validation step |
| 39. Marijnissen 2016 [37] | Ineligible population group | Wrong age group |

| | | |
|---|---|---|
| 40. Boschloo 2014 [38] | Not true multivariable prediction model | Individual risk factor |
| 41. Hofmann 2013 [39] | Review/other design | |
| 42. Taylor 2014 [40] | Outcome not trajectories of dep/anx | |
| 43. Li 2023 [41] | Not true multivariable prediction model | |
| 44. Fatori 2022 [42] | Not true multivariable prediction model | |
| 45. Jacobson 2021 [43] | Outcome not trajectories of dep/anx | |
| 46. Schwartz 2021 [44] | Not true multivariable prediction model | Individual risk factor |
| 47. Scott 2020 [45] | Review/other design | Book chapter |
| 48. Van loo 2020 [46] | Outcome not trajectories of dep/anx | |
| 49. Van Tuijl 2018 [47] | Not true multivariable prediction model | |
| 50. Lee 2017 [48] | Not true multivariable prediction model | |
| 51. Serra-Blasco 2016 [49] | Ineligible population group | Hospital sample |
| 52. Briere 2016 [50] | Not true multivariable prediction model | intervention study |
| 53. Yuka Kudo 2016 [51] | Not true multivariable prediction model | |
| 54. Rei Monden 2016 [52] | Not true multivariable prediction model | |
| 55. Park 2015 [53] | Ineligible population group | Wrong age group |
| 56. Karsten 2013 [54] | Outcome not trajectories of dep/anx | |
| 57. Grant 2013 [55] | Not true multivariable prediction model | |
| 58. Taylor-Clift 2008 [56] | Not true multivariable prediction model | |
| 59. Hahn 2011 [57] | Ineligible population group | hospital sample |
| 60. ten doesschate 2010 [58] | Ineligible population group | hospital sample |
| 61. Karlsson 2008 | Not true multivariable prediction model | and other reasons, hospital sample |
| 62. Hajcack 2025 [59] | Outcome not trajectories of dep/anx | |
| 63. Bansal 2025 [60] | Outcome not trajectories of dep/anx | |
| 64. Remmerswaal 2024 [61] | Outcome not trajectories of dep/anx | |
| 65. Habets 2021 [62] | Outcome not trajectories of dep/anx | |
| 66. Montorsi 2024 [63] | Outcome not trajectories of dep/anx | also old age |
| 67. Pilmeyer 2024 [64] | Outcome not trajectories of dep/anx | |
| 68. Bokma 2020 [65] | Outcome not trajectories of dep/anx | |
| 69. Ray 2021 [66] | Outcome not trajectories of dep/anx | |
| 70. Van Eeden 2021 [67] | Outcome not trajectories of dep/anx | |

| | | |
|---|---|---|
| 71. Librenza-Garcia 2020 [68] | Outcome not trajectories of dep/anx | |
| 72. Lorenzo-Luaces 2020 [69] | Not true multivariable prediction model | |
| 73. Carbonaro 2019 [70] | Not true multivariable prediction model | |
| 74. Struijs 2018 [71] | Not true multivariable prediction model | |
| 75. ten Have 2018 [72] | Not true multivariable prediction model | |
| 76. Spinhoven 2016 [73] | Not true multivariable prediction model | Did not include an internal validation step, individual risk factors |
| 77. Fokkema 2015 [74] | Outcome not trajectories of dep/anx | |
| 78. Hill 2015 [75] | Not true multivariable prediction model | Did not include an internal validation step |
| 79. Wardenaar 2014 [76] | Not true multivariable prediction model | |
| 80. Taylor 2014 [77] | Not true multivariable prediction model | |
| 81. Wardenaar 2012 [78] | Not true multivariable prediction model | |
| 82. Rhebergen 2011 [79] | Not true multivariable prediction model | |
| 83. Sumner 2010 [80] | Review/other design | |
| 84. Batelaan 2010 [81] | Not true multivariable prediction model | |
| 85. Pettit 2009 [82] | Not true multivariable prediction model | |
| 86. Trumpf 2009 [83] | Not true multivariable prediction model | |
| 87. Vuorilehto 2009 [84] | Not true multivariable prediction model | |
| 88. Patton 2008 [85] | Not true multivariable prediction model | |
| 89. Hammen 2008 [86] | Not true multivariable prediction model | |
| 90. Gruenewald 2008 [87] | Not true multivariable prediction model | |
| 91. Lacoviello 2007 [88] | Not true multivariable prediction model | |
| 92. Baldwin 2006 [89] | Not true multivariable prediction model | |
| 93. van den Brink 2001 [90] | Emailed authors for more information - no follow up | |
| 94. Mino 2001 [91] | Ineligible population group | |
| 95. Baldwin 2000 [92] | Ineligible population group | |
| 96. Kivela 2000 [93] | Not true multivariable prediction model | |
| 97. Casten 1999 [94] | Ineligible population group | |
| 98. Katschnig 1998 [95] | Review/other design | |
| 99. Beekman 1995 [96] | Ineligible population group | |
| 100. Scott 1992 [97] | Ineligible population group | |

| | | | |
|---|---|---|---|
| 101. | Warner 1992 [98] | Not true multivariable prediction model | |
| 102. | Grassi 2022 [99] | Ineligible population group | |
| 103. | Scheer 2020 [100] | Outcome not trajectories of dep/anx | |
| 104. | van Tol 2021 [101] | Review/other design | |
| 105. | Michelini 2021 [102] | Not true multivariable prediction model | Risk factors and on internal validation step |
| 106. | Quinlan 2020 [103] | Not true multivariable prediction model | |
| 107. | Cosci 2019 [104] | Review/other design | |
| 108. | Van loo 2018 [105] | Outcome not trajectories of dep/anx | |
| 109. | Bufferd 2018 [106] | Not true multivariable prediction model | |
| 110. | Maarsingh 2018 [107] | Outcome not trajectories of dep/anx | |

### Additional data extraction methods

***Rationale for narrative synthesis approach***

Heterogeneity was expected in several areas including type of prediction model (statistical or machine learning), type/number of predictors included, number of trajectories and method of derivation, demographics of the sample, and depression/anxiety instrument used. Due to this heterogeneity, we planned to report results as a narrative synthesis. We did not specify a minimum number of studies for narrative synthesis. Meta-analysis of an individual prediction model's performance is only recommended when there are multiple (5 or more) external validation studies for the same index model [108].

***Definition of unique prediction model***

We defined a unique prediction model as any model with a unique set of predictors, or different modelling approach (e.g., same predictor set but used a different machine learning approach e.g., gradient boosting vs. random forest). If the same model (same technique and set of predictors) is used to predict facets of a single outcome (e.g., same model used to predict membership of three different trajectories) we count this as a single model (even if a series of binomial regression models are run but using the same method and same set of predictors).

### Additional risk of bias assessment details

#### PROBAST guidance from Moons et al., 2025

***Signalling questions***

Questions may be answered as “Yes”, “Probably Yes”, “No”, “Probably No” or “Unclear”. Moons et al., 2025 [109]- “If the answer to all signalling questions is “Yes” or “Probably Yes,” then risk of bias can be considered low." And “If ≥1 of the answers is “No” or “Probably no,” the judgment could still be “Low risk of bias” but specific reasons should be provided why the risk of bias can be considered low”. Signalling questions are designed to be answered factually to encourage an objective assessment.

***Separate assessments per model***

We have assessed risk of bias based on the primary/best performing model which is reported in the main text. Moons et al., recommend assessing risk of bias individual for each model. We have not done this however, many of the responses to signalling questions will be the same for each model therefore we expect most models reported in the supplement to also have high risk of bias.

***Additional considerations for trajectories***

In the outcome and analysis domains we made additional considerations specific to trajectory outcomes. For example, if a separate prediction was made for membership of each trajectory group,  events-per-variable was calculated using the sample size of the trajectory rather than the overall sample size.

#### EPV calculations

Where sufficient information was given in the paper, we calculated events-per-variable (EPV) for multinomial or binomial logistic regression models in order to support risk of bias assessments in the analysis domain of PROBAST. Papers did not report EPV themselves.

For multinomial models we calculated this by the ratio of the smallest number of observations in the multinomial outcome category divided by the effective number of regression coefficients excluding the intercepts. The number of effective regression coefficients is given by (N of outcome categories – 1)*number of predictors.

For binomial models (one-vs-all, series of binomial models, i.e., binary logistic regression) we defined EPV as the ratio of the number of observations in the smallest of two outcome categories divided by the number of estimated regression coefficients, excluding the intercept.

*Based on 2019 paper by De Jong et al* [110]*.*

*See calculations in tables below.*

#### Supplementary Table 3. EPV calculations for multinomial models

| Author, year | Modelling method and outcome type | EPV / EPVm | N obs in smallest trajectory group | N trajectory groups | N trajectory groups - 1 | N final predictors in best performing model | EPVm |
|---|---|---|---|---|---|---|---|
| **Xiang , 2022** [111] | **Multinomial (categorial outcome)** | **EPVm** | 269 | 4 | 3 | 24 | 3.74 |
| **Schultebraucks 2021** [112] | **Multinomial (categorial outcome)** | **EPVm** | 114 | 4 | 3 | 21 | 1.81 |

#### Supplementary table 4. EPV calculations for binomial one-vs-rest models

| Author, year | Modelling method and outcome type | EPV / EPVm | Trajectory group name | N obs in trajectory group | N in all other trajectory groups together | Smallest of E and F | Parameters= N final predictors in best performing model + 1 | EPV |
|---|---|---|---|---|---|---|---|---|
| **Dinga 2018** [113] | **One-vs-rest (single binomial model; binary outcome)** | **EPV** | **Remitted** | 356 | 448 | 356 | 82 | 4.34 |
| | | | **Improved** | 273 | 531 | 273 | 82 | 3.33 |
| | | | **Chronic** | 175 | 629 | 175 | 82 | 2.13 |
| **Teutenberg 2025** [114] | **One-vs-rest (single binomial model; binary outcome)** | **EPV** | **Remitted** | 178 | 95 | 95 | 28 | 3.39 |
| | | | **Dysthymic** | 30 | 243 | 30 | 28 | 1.07 |
| | | | **Moderate** | 47 | 226 | 47 | 28 | 1.68 |
| | | | **Severe** | 18 | 255 | 18 | 28 | 0.64 |

#### Supplementary table 5. EPV calculations for continuous outcomes

| Author, year | Modelling method (best performing model) and outcome type | EPV (obs per variable, crude) | Trajectory group name | N obs in trajectory group | N in all other trajectory groups together | Smallest of E and F | Parameters= N final predictors in best performing model + 1 | EPV |
|---|---|---|---|---|---|---|---|---|
| **Wardenaar, 2021 [115] - depression** | **Super learner for class probabilities; continuous** | **EPV** | **Chronic** | 1078 | 614 | 614 | 153 | 4.01 |
| | | | **Partial recovery** | 502 | 1190 | 502 | 153 | 3.28 |
| | | | **Full recovery** | 112 | 1580 | 112 | 153 | 0.73 |
| **Wardenaar, 2021 [115] - anxiety** | **Super learner for class probabilities; continuous** | **EPV** | **Full recovery** | 236 | 1457 | 236 | 153 | 1.54 |
| | | | **Partial recovery** | 1306 | 387 | 387 | 153 | 2.53 |
| | | | **Increasing severity** | 151 | 1542 | 151 | 153 | 0.99 |
| **Kessler 2016 [116]** | **Ensemble regression trees for number of years/weeks; continuous** | **EPV** | **Persistent** | not given | n/a | n/a | 14 | Not possible to calculate |
| | | | **Chronic** | 176 | n/a | 176 | 14 | 12.57 |
| **Van Loo 2014 [117]** | **GLM for number of years/weeks; continuous** | **EPV** | **Persistent** | 2869 | 3958 | 2869 | 30 | 95.63 |
| | | | **Chronic** | 3958 | 2869 | 2869 | 30 | 95.63 |
| **Wardenaar 2014 [118]** | **GLM for number of years/weeks; continuous** | **EPV** | **Persistent** | 2869 | 3958 | 2869 | 42 | 68.31 |
| | | | **Chronic** | 3958 | 2869 | 2869 | 42 | 68.31 |
| **Gorham 2022 [119]** | **GLM for number of weeks; continuous** | **EPV** | **n/a** | 92 (overall sample) | n/a | 92 | 11 | 8.36 |

*Note – these are crudely calculated by applying the formula for binomial model EPV to continuous outcomes.*

#### Minor protocol deviations

We pre-registered a protocol for this study on PROSPERO to ensure transparency. We made protocol updates when necessary.

Deviations were minor and included:

- Eligibility criteria clarifications (notes: these were not amendments but clarifications to ensure alignment between reviewers)
  - Clarified definition of prediction model; specified that model should have an internal validation step
  - Samples that included a population based sample along with sample from secondary care would be eligible
- Because there were relatively few full texts for screening, the screening process was unblinded at full text stage and all full-texts were discussed by two reviewers until an agreement on eligibility was made.
- In the protocol we planned to extract secondary data relating to the trajectories themselves (bullets below). We deviated from the protocol by only collecting and reporting the first two items as these were most relevant/required to support our primary outcome of interest (prediction of the trajectories). We were not interested in the details of trajectory modelling.
  - Number and name of trajectories in best fitting model
  - Prevalence of each trajectory/proportion of participants assigned to each class.
  - Average posterior probabilities: measure of classification certainty for individuals within classes
  - Trajectory parameters: the growth factors for each class (e.g., intercept, slope, quadratic terms)

Finally, in the protocol we planned to used PROBAST as a risk of bias assessment tool, however PROBAST-AI was published during our write up [109]. Therefore, we switched to PROBAST-AI.

### Supplementary data extraction

#### Search results and reasons for exclusions

After removing duplicates, we identified 5849 records through database searching and 2800 records through grey literature and citation searches. From these we sought to retrieve 146 full texts for eligibility assessment. We were able to retrieve 126 of these which were screened for eligibility. For one study we contacted the authors but were unable to obtain further information and did not include this study in our review [120].

The most common reason for exclusion was due to studies having the wrong design. More specifically, the majority of the studies falling into this exclusion category were prediction/risk factor studies that did not report the development, validation or performance of a true multivariable prediction model. This was expected because sensitive search terms were used.

Other reasons for exclusion included studies having the wrong outcome (mainly not predicting trajectories of anxiety and depression) or the wrong population (mostly clinical or specialist populations, for example, studies sampling only from hospital populations or studying trajectories of depression after cancer diagnosis). Several studies appeared to use trajectory outcomes (e.g., title or abstract mentioned trajectories or “course” of depression) but when we looked at the full-text outcomes were not in fact trajectories.

#### Interpretation of methods: Kessler 2016, Van Loo and Wardenaar 2014

Van loo 2014 [117] and Wardenaar 2014 [118] are development papers, Kessler 2016 [116] is an external validation paper. The aim of these papers appears to be about whether baseline symptom clusters from the incident episode (+ family history of psychopathology) can predict persistence or chronicity. Wardenaar 2014 also adds predictors to the model (lifetime neuropsychiatric comorbidity) additional to Van Loo 2014. The prediction modelling methods and final predictor lists are not clear. The authors seem to cluster all baseline predictors based on univariable analysis upfront, then from this form multivariable clusters that are used to allocate people into different risk levels i.e., which risk level / cluster of multivariable symptoms is optimal for differentiating/discriminating the outcomes correctly (chronicity and persistence), based on AUC. We deemed these to be multivariable prediction models and eligible for our review.

#### Detailed data extraction tables

*NB: performance measures for models and predictors are listed elsewhere in main text & supplement.*

##### Xiang 2022

111

| | |
|---|---|
| Study details | Country: USA<br>Setting: Population based |
| Original cohort / data source information | Source of data: ABCD study; prospective cohort<br>Original cohort recruitment and sampling method: Mutli-stage probability sample from schools<br>Original recruitment enrollment period: 2016-2018<br>Number of sites: 21<br>Age range of original cohort: 9-10 |

| | |
|---|---|
| Population | Key inclusion and exclusion criteria:<br>• Missing CBCL data<br>• No QC fMRI data<br>• Non-binary gender |
| Baseline characteristics | Mean age (SD): not given<br>Gender (% female): 49%<br>Baseline diagnosis: Healthy |
| Sample size | Total number of participants in baseline sample: 4962<br>N observations missing data: 132 |
| Methods | Type of study (model development or validation): development<br><br>*Prediction model:*<br><br>Number of models reported: 7 models mentioned and reported<br>Model development method: four different multi-class supervised learning for multinomial classification, including K-Nearest Neighbor (KNN), Random Forest (RF), Gradient Boosting Machine (GBM) model and Extreme Gradient Boosting (XGBoost). In supplement also SVM an LDA models.<br>Best performing model (reported in main text): Gradient Boosting Machine (GBM)<br>Internal validation method: 10 fold cross validation<br>External validation: none<br>Handling of missing data: Excluded variables with >10% MD, Multiple imputation of variables with <10% MD<br>Evaluation of clinical utility: none<br><br>*Prediction factors:*<br><br>Selection of candidate predictors: literature based<br>Timing of measurement: Baseline<br>Candidate predictor N: 181<br>Feature selection during modelling: Recursive feature elimination, parameter optimization |

| | |
|---|---|
| | Feature importance method: SHAP ("model-agnostic interpretation method"); represents the deviation from the average predicted value for each case prediction brought by each feature<br>Number of predictors in final model: 24<br>Number of predictors in other models: 24<br>N predictors with missing data: 7 with >10% missingness<br>Blind to outcome: yes, collected at baseline before outcomes<br><br>*Trajectories:*<br><br>Measure of depression/anxiety: CBCL<br>Blind to predictors: probably not, parent reported and parents would have knowledge of many predictors<br>Method: LCGA<br>Data type: categorical, groups<br>Repeated measures & years of follow up: 3 timepoints over 2 year follow up<br>Trajectory group or outcome defined (including sample size):<br>Decreasing (n=433)<br>Persistently high (n=269)<br>Increasing (n=536)<br>Persistently low (n=3724) |

#### Wardenaar 2021

115

| | |
|---|---|
| Study details | Country: Netherlands<br>Setting: Population, primary and secondary care |
| Original cohort / data source information [121] | Source of data: NESDA; prospective cohort<br>Original cohort recruitment and sampling method: Multistage stratified in community, primary and secondary care<br>Original recruitment enrollment period: 2004-2006<br>Number of sites: 65 GP practices, 17 mental health institutions<br>Age range of original cohort: 18-65 |
| Population | Key inclusion and exclusion criteria: |

| | |
|---|---|
| | Exclusion criteria at baseline were: not being fluent in Dutch, a primary diagnosis of a psychotic, obsessive-compulsive disorder, bipolar disorder or severe addiction disorder. |
| Baseline characteristics | Mean age (SD): 41.3 (±12.4) years (note, in background section 41.6 years is quoted. 41.3 taken from Table 1)<br>Gender (% female): 66%<br>Baseline diagnosis: diagnosis of dysthymia, major depressive disorder (MDD) or an anxiety disorder within 6 months before baseline |
| Sample size | Total number of participants in baseline sample: 1693<br>N observations missing data: Overall, only 1.7% of the data were missing |
| Methods | *General:*<br><br>Type of study (model development or validation): development<br><br>*Prediction model:*<br><br>Number of models reported: 8 models mentioned, 7 reported. 6 base learner models, 1 “super learner” and OLS regression model (results for the regression model not reported).<br>Model development method: Elasticnet, Random Forest with trees hyperparameter set to 100 200 and 500, GBM, SVM, Super learner<br>Best performing model (reported in main text): super learner<br>Internal validation method: 10-fold cross validation<br>External validation: none<br>Handling of missing data: Multiple imputation; PMM<br>Evaluation of clinical utility: none<br><br>*Prediction factors:*<br><br>Selection of candidate predictors: all available in dataset<br>Timing of measurement: baseline<br>Candidate predictor N: 152<br>Feature selection during modelling: unclear, looks like all 152 used and 15 top predictors in an OLS regression model but model not reported |

| | |
|---|---|
| | Feature importance: For each outcome, the importance of each determinant in the SL was investigated by evaluating the change in the MSESL (i.e. MSE difference) when the given determinant's data were randomly scrambled. Spearman correlations between each determinant and the model-predicted score were calculated to gain insight into the determinants' roles in the SL.<br>Number of predictors in final model: 152<br>Number of predictors in other models: 152 in all<br>N predictors with missing data: n=66 predictors had 1 or more missing value (range: 5.9%-15.8%)<br>Blind to outcome: yes, collected at baseline before outcome<br><br>*Trajectories:*<br><br>Measure of depression/anxiety: IDS-SR for depression and BAI for anxiety<br>Method: LCGA<br>Blind to predictors: self-report so likely not<br>Data type: categorical, groups<br>Repeated measures and follow-up: 5 timepoints over 9 year follow up<br>Trajectory group or outcome defined (including sample size):<br>Depression, chronic (n=1078)<br>Depression, partial recovery (n=502)<br>Depression, full recovery (n=112)<br>Anxiety, full recovery (n=236)<br>Anxiety, partial recovery (n=1306)<br>Anxiety, increasing severity (n=151) |

#### Dinga 2018

113

| | |
|---|---|
| Study details | Country: Netherlands<br>Setting: Population, primary and secondary care |

| | |
|---|---|
| Original cohort / data source information [121] | Source of data: NESDA; prospective cohort<br>Original cohort recruitment and sampling method: Multistage stratified in community, primary and secondary care<br>Original recruitment enrollment period: 2004-2006<br>Number of sites: 65 GP practices, 17 mental health institutions<br>Age range of original cohort: 18-65 |
| Population | Key inclusion and exclusion criteria:<br><br>NESDA: not being fluent in Dutch, a primary diagnosis of a psychotic, obsessive-compulsive disorder, bipolar disorder or severe addiction disorder.<br><br>Additional study specific selection criteria: presence of a DSM-IV MDD or dysthymia diagnosis (or both) in the past 6 months at baseline, established using the structured Composite International Diagnostic Interview (CIDI, version 2.1); confirmation of depressive symptoms in the month prior to baseline either by the CIDI or the Life Chart Interview (LCI); and availability of 2-year follow-up data on DSM-IV diagnosis and depressive symptoms measured with the LCI. |
| Baseline characteristics | Mean age (SD): 41.9 (±12.2) years<br>Gender (% female): 65%<br>Baseline diagnosis: DSM-IV MDD or dysthymia diagnosis (or both) in the past 6 months at baseline |
| Sample size | Total number of participants in baseline sample: 804<br>N observations missing data: unclear |
| Methods | *General:*<br><br>Type of study (model development or validation): development<br><br>*Prediction model:*<br>Number of models reported: 4 models mentioned and reported<br>Model development method: Multinomial classification and one-vs-all discrimination using: generalization of penalized (elastic-net) logistic regression<br>Best performing model (reported in main text): full model combining all predictors<br>Internal validation method: 10-fold cross validation<br>External validation: none |

| | |
|---|---|
| | Handling of missing data: multiple imputation<br>Evaluation of clinical utility: none<br><br>*Prediction factors:*<br><br>Selection of candidate predictors: literature based<br>Timing of measurement: baseline<br>Candidate predictor N: 81<br>Feature selection during modelling: stability selection - stability paths indicating how often each variable in the model is selected as a function of the regularization applied.<br>Feature importance: not specifically mentioned<br>Number of predictors in final model: full model=81<br>Number of predictors in other models: biological model=18, clinical model=55, personality model=5<br>N predictors with missing data: 25/80, Of those variables, the median number of missing values per variable was 11 (1.4% of the sample). 23 out of 25 missing variables contained less than 7% of missing values with an exception of cortisol ROCg and cortisol ROCi with 38% of missing values.<br>Blind to outcome: yes, collected at baseline<br><br>*Trajectories:*<br><br>Measure of depression/anxiety: CIDI<br>Method: LCGA<br>Blind to predictors: by trained assessors but no mention of blinding<br>Data type: categorical, groups<br>Repeated measures and follow up: 24 timepoint over 2 year follow up<br>Trajectory group or outcome defined (including sample size):<br>Remitted (n=356)<br>Improved (n=273)<br>Chronic (n=175) |

#### Kessler 2016

116

| | |
|---|---|
| Study details | Country: USA<br>Setting: population based |
| Original cohort / data source information | Source of data: US National Comorbidity survey[122] 1 + 2; longitudinal follow up of sub-sample from cross-sectional survey<br>Original cohort recruitment and sampling method: multi-stage, clustered area probability sampling of non-institutionalized U.S. households<br>Original recruitment enrollment period: 2001-2003<br>Number of sites: 62 primary sampling units (PSUs – which were geographical areas), each PSU into segments of between 50 and 100 housing units. Response rate above 70% among primary and 80.4% among secondary predesignated respondents<br>Age range of original cohort: 15-54 |
| Population | Key inclusion and exclusion criteria:<br><br>Original survey: English-speaking adults ages 18 or older living in the non-institutionalized civilian household population of the coterminous US (excluding Alaska and Hawaii) plus students living in campus group housing who have a permanent household address |
| Baseline characteristics | Mean age (SD): unclear<br>Gender (% female): 50%<br>Baseline diagnosis: Lifetime MDD |
| Sample size | Total number of participants in baseline sample:<br>N observations missing data: unclear |
| Methods | *General:*<br><br>Type of study (model development or validation): external validation in an independent household sample using |

| | |
|---|---|
| | machine learning methods; however, not true external validation as predictor sets appear to be different to development studies and not carried out by independent authors.<br><br>*Prediction model:*<br><br>Number of models reported: 2 mentioned, 1 reported<br>Model development method: Ensemble regression trees ("machine learning" validation of regression models developed in other papers), a logistic regression model mentioned but not reported<br>Best performing model (reported in main text): Ensemble regression trees<br>Internal validation method: 10-fold cross validation<br>Handling of missing data: multiple imputation<br>Evaluation of clinical utility: none<br><br>*Prediction factors:*<br><br>Selection of candidate predictors: regression models in previous studies<br>Timing of measurement: baseline<br>Candidate predictor N: 23 (although unclear)<br>Feature selection during modelling: unclear<br>Feature importance: not mentioned<br>Number of predictors in final model: "between 9-13" in machine learning model<br>Number of predictors in other models: 23 in statistical model<br>N predictors with missing data: unclear<br>Blind to outcome: no, some predictors taken from survey 2 (i.e., not baseline) therefore outcome data known at this point<br><br>*Trajectories:*<br><br>Measure of depression/anxiety: CIDI<br>Blind to predictors: collected retrospectively and self-reported so participants would be aware of own predictor info (not blinded).<br>Method: Proportion of years since age of onset where subject had MDD over 10-12 year follow up |

| | |
|---|---|
| | Data type: continuous<br>Repeated measures: n/a<br>Years of follow up: 10-12 years<br>Trajectory group or outcome defined (including sample size):<br>Persistent (years with episode 2+ weeks) (n=not given)<br>Chronic (lasting most days in year) (n=176)<br>*Mean (se) number of years in episode = 2.0 (0.2), 90th percentile = 9 years. |

#### Van Loo 2014

117

| | |
|---|---|
| Study details | Sponsorship source:<br>Country: 16 countries<br>Setting: population based |
| Original cohort / data source information | Source of data: WMH Surveys[123]; retrospective cohort<br>Original cohort recruitment and sampling method: “Most WMH surveys are based on stratified multistage clustered area probability household samples in which samples of areas equivalent to counties or municipalities in the US were selected in the first stage followed by one or more subsequent stages of geographic sampling (e.g., towns within counties, blocks within towns, households within blocks) to arrive at a sample of households, in each of which a listing of household members was created and one or two people were selected from this listing to be interviewed. No substitution was allowed when the originally sampled household resident could not be interviewed. These household samples were selected from Census area data in all countries other than France (where telephone directories were used to select households) and the Netherlands (where postal registries were used to select households). Several WMH surveys (Belgium, Germany, Italy, Poland, Poland 2, Spain-Murcia) used municipal resident registries to select respondents without listing households. The Japanese sample is the only totally un-clustered sample, with households randomly selected in each of the 11 metropolitan areas and one random respondent selected in each sample household. The sample for the Qatar survey was drawn from a national list of cellular telephone numbers.” The average weighted response rate was 73.7% (range: 55.1-95.2%).<br>Original recruitment enrollment period: 2001-2009<br>Number of sites: unclear – see above sampling description<br>Age range of original cohort: 18-100 years |

| | |
|---|---|
| Population | Key inclusion and exclusion criteria: adults aged 18+ with lifetime DSM-IV/CIDI MDD. Other inc/exc not clear. |
| Baseline characteristics | Mean age (SD): not given<br>Gender (% female): not given<br>Baseline diagnosis: lifetime DSM-IV/CIDI MDD |
| Sample size | Total number of participants in baseline sample: 8261<br>N observations missing data: unclear |
| Methods | *General:*<br><br>Type of study (model development or validation): development<br><br>*Prediction model:*<br><br>Number of models reported: unclear, possibly 6 cluster models ("3 through to 8 clusters")<br>Model development method: GLM with Lasso penalised regression<br>Best performing model (reported in main text): 3 cluster model – not clear<br>Internal validation method: Bootstrapping and cross-validation for initial clustering step - not mentioned for final model, general reporting of methods is not clear.<br>External validation: none<br>Handling of missing data: not clear<br>Evaluation of clinical utility: none<br><br>*Prediction factors:*<br><br>Selection of candidate predictors: available in dataset, focus on clinical/depression subtype characteristics of baseline and previous episodes<br>Timing of measurement: baseline<br>Candidate predictor N: ~35, not clear<br>Feature selection during modelling: based on clustering to generate risk clusters that predict trajectories<br>Feature importance: not mentioned<br>Number of predictors in final model: unclear, counted up to 29<br>Number of predictors in other models: n/a<br>N predictors with missing data: unclear |

| | |
|---|---|
| | Blind to outcome: retrospective; subject to recall bias which may be different between participants and means outcome/predictor info collected at same time. Unclear whether blinded.<br><br>*Trajectories:*<br><br>Measure of depression/anxiety: CIDI<br>Blind to predictors: outcome/predictor info collected at same time. Unclear if blinded.<br>Method: Proportion of years since age of onset where subject had MDD over 10-12 year follow up<br>Data type: continuous<br>Repeated measures: n/a<br>Years of follow up:<br>Trajectory group or outcome defined (including sample size):<br>Persistent (years with episode 2+ weeks) (n=2869)<br>Chronic (lasting most days in year) (n=3958) |

#### Wardenaar 2014

119

| | |
|---|---|
| Study details | Sponsorship source:<br>Country: 16 countries<br>Setting: population based |
| Original cohort / data source information | Source of data: WMH Surveys [123]; retrospective cohort<br>Original cohort recruitment and sampling method: “Most WMH surveys are based on stratified multistage clustered area probability household samples in which samples of areas equivalent to counties or municipalities in the US were selected in the first stage followed by one or more subsequent stages of geographic sampling (e.g., towns within counties, blocks within towns, households within blocks) to arrive at a sample of households, in each of which a listing of household members was created and one or two people were selected from this listing to be interviewed. No substitution was allowed when the originally sampled household resident could not be interviewed. These household samples were selected from Census area data in all countries other than France (where telephone directories were used to select households) and the Netherlands (where postal registries were used to select |

| | |
|---|---|
| | households). Several WMH surveys (Belgium, Germany, Italy, Poland, Poland 2, Spain-Murcia) used municipal resident registries to select respondents without listing households. The Japanese sample is the only totally un-clustered sample, with households randomly selected in each of the 11 metropolitan areas and one random respondent selected in each sample household. The sample for the Qatar survey was drawn from a national list of cellular telephone numbers." The average weighted response rate was 73.7% (range: 55.1-95.2%).<br>Original recruitment enrollment period: 2001-2009<br>Number of sites: unclear – see above sampling description<br>Age range of original cohort: 18-100 years |
| Population | Key inclusion and exclusion criteria: adults aged 18+ with lifetime DSM-IV/CIDI MDD. Other inc/exc not clear. |
| Baseline characteristics | Mean age (SD): not given<br>Gender (% female): not given<br>Baseline diagnosis: lifetime DSM-IV/CIDI MDD |
| Sample size | Total number of participants in baseline sample: 8261<br>N observations missing data: unclear |
| Methods | *General:*<br><br>Type of study (model development or validation): development<br><br>*Prediction model:*<br><br>Number of models reported: not clear, just 1<br>Model development method: Classification trees with recursive partitioning, followed by penalised regression methods (lasso, ridge penalty, elastic net penalty)<br>Best performing model (reported in main text):3 cluster model<br>Internal validation method: Bootstrapping and cross-validation for initial clustering step - not mentioned for final model, general reporting of methods is not clear.<br>External validation: none<br>Handling of missing data: not clear<br>Evaluation of clinical utility: none |

| | |
|---|---|
| | *Prediction factors:*<br><br>Selection of candidate predictors: available in dataset, focus on clinical/depression subtype characteristics of baseline and previous episodes<br>Timing of measurement: baseline<br>Candidate predictor N: ~45, not clear<br>Feature selection during modelling: based on clustering to generate risk clusters that predict trajectories<br>Feature importance: not mentioned<br>Number of predictors in final model: unclear, mentions, 22 retained after LASSO.<br>N predictors with missing data: unclear<br>Blind to outcome: retrospective; subject to recall bias which may be different between participants and means outcome/predictor info collected at same time, unclear if blinded<br><br>*Trajectories:*<br><br>Measure of depression/anxiety: CIDI<br>Blind to predictors: outcome/predictor info collected at same time, unclear if blinded<br>Method: Proportion of years since age of onset where subject had MDD over 10-12 year follow up<br>Data type: continuous<br>Repeated measures: n/a<br>Years of follow up:<br>Trajectory group or outcome defined (including sample size):<br>Persistent (years with episode 2+ weeks) (n=2869)<br>Chronic (lasting most days in year) (n=3958) |

#### Gorham 2022

119

| | |
|---|---|
| Study details | Sponsorship source:<br>Country: USA<br>Setting: population based, primary and secondary care |

| | |
|---|---|
| Original cohort / data source information | Source of data: NIMH CAT-D longitudinal study; Prospective cohort<br>Original cohort recruitment and sampling method: Participants are recruited locally and nationwide via mailings to selected physicians, announcements in newsletters, and contacts with support groups and approved websites.<br>Original recruitment enrollment period: since 2017<br>Number of sites: no information<br>Age range of original cohort: 11-17 |
| Population | Key inclusion and exclusion criteria:<br><br>Original CAT-D study:<br>To be included as an MDD participant in this characterization study, one must be between the ages of 11-17 at the time of enrolment and must have a current diagnosis of MDD via the DSM-5 within the last six months. To be included as an s-MDD (subthreshold depression) participant in this characterization study, one must be between the ages of 11-17 at the time of enrolment, must have an episode of depressed mood or loss of interest or pleasure lasting at least one week, and must have at least two of the seven other DSM-5 associated symptoms for major depression occurring in the last six months. To be included as a healthy volunteer (HV) participant in this characterization study, one must be between the ages of 11-17 at the time of enrolment, must be competent to assent (and parents must be competent to consent), must be willing to participate in research and be willing to undergo psychiatric interviews, must speak English, and must have a primary care clinician in the community.<br><br>Exclusionary criteria for all participants include: a diagnosis of schizophrenia, schizophreniform disorder, schizoaffective illness, bipolar disorder, severe Autism Spectrum Disorder, Anorexia Nervosa, or other severe eating disorders; an IQ of <70; depressive symptoms that are due to effects of drugs of abuse or a neurological condition; a diagnosis of alcohol or other substance use disorders; current active suicidal ideation; repeated self-harm in the context of interpersonal conflict; having an immediate family member who works at the NIMH; and a serious medical condition such as epilepsy or heart disease. |
| Baseline characteristics | Mean age (SD): 15.8 (±1.3) years<br>Gender (% female): 72% female<br>Baseline diagnosis: Healthy, subthreshold or full depression diagnosis (see definitions above) |
| Sample size | Total number of participants in baseline sample: 92<br>N observations missing data: n=13 |

| | |
|---|---|
| Methods | *General:*<br><br>Type of study (model development or validation): development<br><br>*Prediction model:*<br><br>Number of models reported: 11<br>Model development method: regression (no further information) – this is a borderline prediction model study; appears to be more interested in a single prediction factor (family history) however, meets our criteria; several multivariable models included with an internal validation step<br>Best performing model (reported in main text): family history, MFQ and CASE<br>Internal validation method: 8-fold cross validation<br>External validation: none<br>Handling of missing data: multiple imputation<br>Evaluation of clinical utility: none<br><br>*Prediction factors:*<br><br>Selection of candidate predictors: unclear<br>Timing of measurement: baseline (mostly)<br>Candidate predictor N: 11<br>Feature selection during modelling: unclear<br>Feature importance: not mentioned<br>Number of predictors in final model: up to 11<br>Number of predictors in other models: null model=8, others 9<br>N predictors with missing data: unclear<br>Blind to outcome: all collected at baseline apart from CASE<br><br>*Trajectories:*<br><br>Measure of depression/anxiety: Kiddy-SADS-PL DSM 5 depression screener and supplement<br>Method: Number of weeks spent in depressive episode from baseline to 1 year follow up<br>Blind to predictors: unclear |

| | |
|---|---|
| | Data type: continuous<br>Repeated measures: n/a<br>Years of follow up: 1 year<br>Trajectory group or outcome defined (including sample size): n=92 |

#### Schultebraucks 2021

112

| | |
|---|---|
| Study details | Sponsorship source:<br>Country: USA<br>Setting: population based |
| Original cohort / data source information | Source of data: Health and Retirement study [124], household sample<br>Original cohort recruitment and sampling method: Area-Based Multistage Probability Sampling. Oversample of Blacks, Hispanics (primarily Mexican Americans), and Florida residents was drawn to increase the sample size of Blacks and Hispanics as well as those who reside in the state of Florida. Weighting applied.<br>Original recruitment enrollment period: 1992<br>Number of sites: unclear<br>Age range of original cohort: 51-61 years |
| Population | Key inclusion and exclusion criteria:<br><br>Original HRS study: adults residing in households (i.e., community dwelling, non-institutionalized individuals) in the contiguous United States born between 1931 and 1941 (i.e., those who were between the ages of 51–61 in 1992 |
| Baseline characteristics | Mean age (SD): 55.9 (±8.5) years<br>Gender (% female): 64%<br>Baseline diagnosis: healthy but has experienced 1 major life stressor from following list: bereavement, myocardial infarction, divorce, cancer, or job loss. |
| Sample size | Total number of participants in baseline sample: 2071<br>N observations missing data: 3% |

| | |
|---|---|
| Methods | *General:*<br><br>Type of study (model development or validation): development<br><br>*Prediction model:*<br><br>Number of models reported: 3 models mention, results only given for 1<br>Model development method: supervised learning for multinomial classification using a multilayer feedforward neural network to train a deep neural net (DNN). Logistic regression (Benchmark models mentioned in supplement but no results).<br>Best performing model (reported in main text): neural network<br>Internal validation method: 10-fold cross validation<br>External validation: none<br>Handling of missing data: complete case analysis? - "problematic missingness (>2%) for polygenic risk scores" – individuals filtered out by QC process. For variables with more than 35% missingness were removed.<br>Evaluation of clinical utility: none – however note that applied LIME to make results more interpretable.<br><br>*Prediction factors:*<br><br>Selection of candidate predictors: literature based<br>Timing of measurement: baseline (polygenic risk scores)<br>Candidate predictor N: 21<br>Feature selection during modelling:<br>Feature importance: LIME "to estimate which set of features, on average, influenced the classification of depressive symptom trajectories the most"<br>Number of predictors in final model: 21<br>Number of predictors in other models: unclear<br>N predictors with missing data: problematic missingness (>2%) for polygenic risk scores<br>Blind to outcome: yes – PGS measured at baseline – "genome" objective and blind to outcome.<br><br>*Trajectories:* |

| | |
|---|---|
| | Measure of depression/anxiety: CES-D<br>Blind to predictors: unclear – probably yes – self report, participants will be blinded to genome but not phenotype<br>Method: LGMM<br>Data type: categorical, groups<br>Repeated measures and follow up: 3 timepoints over 2 year follow up<br>Trajectory group or outcome defined (including sample size):<br>Resilience (n=1638)<br>Recovery (n=160)<br>Emerging depression (n=159)<br>Pre-existing and chronic depression (n=114) |

#### Teutenberg 2025

114

| | |
|---|---|
| Study details | Sponsorship source:<br>Country: Germany<br>Setting: Population, primary and secondary care |
| Original cohort / data source information | Source of data: MACS (Marburg-Münster Affective Disorders Cohort Study, http://for2107.de/)<br>Original cohort recruitment and sampling method:" Recruitment of patients took place via departments of the university and surrounding psychiatric hospitals of Marburg and Münster, Germany, doctors' surgeries, newspaper advertisements, and flyers."<br>Original recruitment enrollment period: unclear<br>Number of sites: 2 center research consortium; 13 sites listed in website<br>Age range of original cohort: 18-65 |
| Population | Key inclusion and exclusion criteria:<br><br>MACS: Patients were excluded from participation in the study if they were diagnosed with any life-time substance dependence disorder, had a history of severe neurological or medical disorders, did not have a West-European ancestry, or were currently using benzodiazepines. Participants of non-West-European ancestry were excluded because the FOR2107 MACS cohort mainly focuses on genetic and neuroimaging studies, aiming for more genetic |

| | |
|---|---|
| | homogeneity. To be included in the current analyses, patients furthermore had to provide available follow-up and life chart data |
| Baseline characteristics | Mean age (SD): 36.7 (±13.2) years<br>Gender (% female): 67%<br>Baseline diagnosis: patients and healthy volunteers |
| Sample size | Total number of participants in baseline sample: 571<br>N observations missing data: n=14– "Before computing predictive models, data was inspected for missing values, however, neither subjects nor features exceeded missing values >5%, therefore, no data had to be discarded." |
| Methods | *General:*<br><br>Type of study (model development or validation): development<br><br>*Prediction model:*<br><br>Number of models reported: 3 models<br>Model development method: Random forest classifiers<br>Best performing model (reported in main text): explorative<br>Internal validation method: cross-validation<br>External validation: none<br>Handling of missing data: Existing missing data was imputed using k-nearest neighbor imputation implemented in a pipeline, avoiding information leakage.<br>Evaluation of clinical utility: none<br><br>*Prediction factors:*<br><br>Selection of candidate predictors: literature based<br>Timing of measurement: baseline<br>Candidate predictor N: 27<br>Feature selection during modelling: "Hyperparameters were tuned in the inner folds via randomized grid search, assessing the best model across 50 parameter combinations based on mean balanced accuracy (BACC)."<br>Feature importance: "RFCs were a suitable classifier as they facilitate interpretability of feature importance"<br>Number of predictors in final model: explorative model=27 |

| | |
|---|---|
| | Number of predictors in other models: good evidence predictor model=7, strong evidence predictor model=3<br>N predictors with missing data: not clear<br>Blind to outcome: collected at baseline, yes<br><br>*Trajectories/outcome:*<br><br>Measure of depression/anxiety: SCID-I<br>Blind to predictors: trained interviewers, no mention of blinding<br>Method: Model-based clustering approach and latent profile analysis<br>Data type: categorical, groups<br>Repeated measures and follow up: 24 timepoints; reported retrospectively by life charting method for each month over 2 year interval<br>Trajectory group or outcome defined (including sample size):<br>Remitted (n=178)<br>Dysthymic (n=30)<br>Moderate (n=47)<br>Severe (n=18) |

#### Supplementary table 6. Number of unique models in each study

| Author, Year | UNIQUE MODELS MENTIONED RELEVANT TO OUTCOMES FOR REVIEW | UNIQUE MODELS FOR WHICH PERFORMANCE REPORTED | WHICH MODEL(S) REPORTED IN MAIN TEXT (optimal performance) | OTHER REPORTED MODELS |
|---|---|---|---|---|
| **Xiang 2022** | 7 | 7 | GBM | 4 additional models in main text and 3 models in supplement |
| **Wardenaar 2021** | 8 | 7 | Super learner | 6 base learners<br>OLS regression model mentioned but results reported |
| **Dinga 2018** | 4 | 4 | Full model | 3 additional models in main text (clincal model, personality trait model, biological model) |
| **Kessler 2016** | 2 | 1 | Ensemble regression tree model | 1 additional logistic regression model |
| **Van Loo 2014** | 6 | 2 | 3 cluster model ?- unclear | Methods unclear ("3 through 8 cluster solutions") |
| **Wardenaar 2014** | 1 | 1 | 3 cluster model ?- unclear | Methods unclear (only the 3 cluster solution from Van Loo 2014) |
| **Gorham 2022** | 11 | 11 | FH+MFQ+CASE+Null | 4 additional models in main text (null model, FH model, MFQ model, MFQ+FH+null)<br>6 additional models in supplement |
| **Schultebraucks 2021** | 3 | 1 | Neural network | 2 additional "benchmark" models in supplement - 1 sociodemographic model, 1 with only PGS for depression, no results given |
| **Teutenberg 2025** | 3 | 3 | Explorative model | 2 additional models: Strong evidence predictors, good evidence predictors |
| **Total** | **45** | **37** | | |

### Supplementary table 7. Performance statistics of other models

| Author, Year | Model name | N of predictors used | Performance statistics |
|---|---|---|---|
| Xiang 2022 | XGBOOST | 24 | Macro-average AUC=0.78 |
| | Random forest | 24 | Macro-average AUC=0.76 |
| | KNN | 24 | Macro-average AUC=0.64 |
| | SVM with linear kernel | 24 | Macro-average AUC=0.75 |
| | SVM with radial basis function kernel | 24 | Macro-average AUC=0.74 |
| | LDA | 24 | Macro-average AUC=0.75 |
| Wardenaar 2021 | Elastic net | 152 | Depression chronic course: MSE(SE)=0.13 (0.003)<br>Depression partial recovery: MSE(SE)=0.11 (0.003)<br>Depression full recovery: MSE(SE)=0.05 (0.004)<br>Anxiety full recovery: MSE(SE)= 0.10 (0.005)<br>Anxiety partial recovery: MSE(SE)=0.12(0.004)<br>Anxiety increasing severity: MSE(SE)=0.06 (0.004) |
| | Random forest (100, 250, 500) - three models, MSE(SE) same for all three | 152 | Depression chronic course: MSE(SE)=0.14 (0.003)<br>Depression partial recovery: MSE(SE)=0.12 (0.003)<br>Depression full recovery: MSE(SE)= 0.05 (0.004)<br>Anxiety full recovery: MSE(SE)= 0.10 (0.004)<br>Anxiety partial recovery: MSE(SE)=0.12 (0.004)<br>Anxiety increasing severity: MSE(SE)= 0.06 (0.004) |
| | Gradient boosting | 152 | Depression chronic course: MSE(SE)=0.13 (0.003)<br>Depression partial recovery: MSE(SE)=0.12 (0.003)<br>Depression full recovery: MSE(SE)=0.05 (0.004)<br>Anxiety full recovery: MSE(SE)= 0.10 (0.005)<br>Anxiety partial recovery: MSE(SE)=0.12 (0.004)<br>Anxiety increasing severity: MSE(SE)=0.06 (0.004) |

| | | | |
|---|---|---|---|
| | SVM | 152 | Depression chronic course: MSE(SE)=0.14 (0.004)<br>Depression partial recovery: MSE(SE)=0.12 (0.004)<br>Depression full recovery: MSE(SE)=0.05 (0.005)<br>Anxiety full recovery: MSE(SE)=0.10 (0.006)<br>Anxiety partial recovery: MSE(SE)=0.13 (0.005)<br>Anxiety increasing severity: MSE(SE)=0.06(0.005) |
| **Dinga 2018** | Biological model | 18 | Remitted; AUROC = 0.57<br>Improved; AUROC = 0.56<br>Chronic; AUROC = 0.61 |
| | Clinical model | 55 | Remitted; AUROC = 0.68<br>Improved; AUROC = 0.59<br>Chronic; AUROC = 0.64 |
| | Personality traits | 5 | Remitted; AUROC = 0.63<br>Improved; AUROC = 0.58<br>Chronic; AUROC = 0.57 |
| **Teutenberg 2025** | Strong evidence predictor set | 4 | Remitted; AUROC = 59.31 (0.55)<br>Dysthymic; AUROC = 55.84 (1.84)<br>Moderate; AUROC = 55.87 (0.92)<br>Severe; AUROC = 66.18 (5.55) |
| | Good evidence predictor set | 7 | Remitted; AUROC = 71.32 (0.64)<br>Dysthymic; AUROC = 50.87 (5.75)<br>Moderate; AUROC = 64.48 (2.29)<br>Severe; AUROC = 61.67 (1.26) |
| **Kessler 2016** | Logistic regression model | 23 | Persistent AUC = 0.68<br>Chronic, AUC = 0.62 |
| **Gorham 2022** | null model | 8 | RMSE (in weeks), bootstrap 99.9% CIs - 18.06 (15 - 24) |
| | family history model | 9 | RMSE (in weeks), bootstrap 99.9% CIs - 15.72 (13 - 24) |
| | MFQ model | 9 | RMSE (in weeks), bootstrap 99.9% CIs - 16.5 (14.5 - 21.5) |
| | FH+MFQ model | 10 | RMSE (in weeks), bootstrap 99.9% CIs - 17.5 (not given) |

### Detailed risk of bias assessment for best performing models

#### Completed PROBAST-AI tables for best performing model:

*Notes: applicability refers to concern that model does not match the review question or the assessor's intended use for the relevant domain.

##### Xiang 2022.

| **Publication reference** | **Xiang 2022** |
|---|---|
| **Model of interest (being evaluated)** | **GBM model** |
| **Domain 1 - Participants** | **Domain rating Y (yes) / PY (probably yes) / PN (probably no) / N (no) / NI (no information)** |
| Describe the sources of data and criteria for participant selection: Prospective longitudinal cohort study. Sampling methods used to ensure nationally representative (although some differences between ABCD parent sample and study sample). Mutli-stage probability sample from multiple schools (21 sites). Key exclusion criteria in study sample included: non-binary gender, missing CBCL (outcome) data and fMRI data that was not QC'd (samples for second 2 exclusions were were large and may have introduced selection bias – however, no information comparing to original sample). | |
| Were appropriate data sources used, e.g., cohort, randomized controlled trial, or nested case-control study data? | Y |
| Was an appropriate study design used? | Y |
| Did the in- and exclusions of study participants result in a representative dataset? | NI |
| DOMAIN 1 - OVERALL QUALITY CONCERN | LOW |
| DOMAIN 1 - APPLICABILITY | LOW |
| Domain 2 - Predictors | |
| List and describe predictors included in the final prediction model, how they were defined and assessed, and their timing of assessment: predictors collected at baseline. Validated scales used. No indication that they were defined/assessed or processed differently across different participants (no info). Possible issue - no real world setting where academic + fMRI data both available at time of assessment. | |
| Were predictors defined and assessed in a similar way for all participants? | NI |
| Was any pre-processing of predictors similar for all participants? | NI |
| Were predictor assessments made without knowledge of outcome data? | Y |
| Are all predictors available at the time the model is intended to be used? | PN |

| | |
|---|---|
| DOMAIN 2 - OVERALL QUALITY CONCERN | HIGH |
| DOMAIN 2 - APPLICABILITY | LOW |
| Domain 3 - Outcome | |
| Describe the outcome, how it was defined and determined, and the time interval between predictor assessment and outcome determination: repeated CBCL measures. CBCL is validated in this age group. However, parent reported (introduces some bias and possibly variability between participants). Also parents would be aware of some predictors when determining outcome (not blind). Longer timeframe (outcome may not have manifested yet) and more repeated measures would have improved outcome measure. | |
| Were outcomes defined and assessed appropriately? | PY |
| Was the outcome defined and determined in a similar way for all participants? | PY |
| Was the outcome determined without knowledge of predictor information? | PN |
| Was the time interval between predictor assessment and outcome determination appropriate? | PN |
| DOMAIN 3 - OVERALL QUALITY CONCERN | HIGH |
| DOMAIN 3 - APPLICABILITY | LOW |
| Domain 4 - Analysis | |
| Describe the numbers of participants, number of candidate predictors, number of outcome events. Describe how the prediction model was developed (e.g., with respect to modelling technique, predictor selection, and classification or risk group definition). Describe the performance measures of the prediction model, e.g., (re)calibration, discrimination, (re)classification, net benefit, and whether they were adjusted for optimism:<br>The authors used a multinomial classification model. EPV = 3.74 based on n=269 observations in smallest trajectory group and 24 predictors in the final / best performing model. The overall sample size was n=4962. Continuous variables were scaled to the range of [0,1] using the min-max method. To address the class imbalance problem, we applied<br>the Synthetic Minority Over-sampling Technique (SMOTE). Predictors were selected using recursive feature elimination; parameters then optimized by train function in R and cross-validation. Discrimination (AUC) is reported but no calibration metrics (are calibration metrics relevant for classification models?). Classification measures are reported.<br><br>Describe missing data on predictors and outcomes as well as methods used for handling<br>these missing data:<br>n=132 observations missing. Unclear how many predictors had missing data? Excluded variables with >10% MD. Multiple imputation of variables with <10% MD. Imputation is good but 10% threshold quite high. | |
| Was model evaluation based on only apparent performance (i.e., training dataset only) avoided? | Y |
| Was there evidence that the sample size was reasonable? (development and evaluation samples) | N |
| Were continuous and categorical predictors handled appropriately? | PY |
| Were participants with missing data handled appropriately? | PY |
| If methods to address class imbalance were used, was the model or the model predictions recalibrated? | NI |
| If resampling methods were used to evaluate model performance, were all model development steps replicated in the resampling process? | NI |

| | |
|---|---|
| If data splitting was done to create training and test datasets, was there evidence that data leakage was avoided? | NI |
| Was the predictive performance of the model evaluated appropriately, e.g., calibration, discrimination, and net benefit? | PN |
| Were methods used to address potential model overfitting? | Y |
| DOMAIN 4 - OVERALL QUALITY CONCERN | HIGH |

#### Wardenaar 2021

| **Publication reference** | **Wardenaar 2021** |
|---|---|
| **Model of interest (being evaluated)** | **Super Learner Model** |
| Domain 1 - Participants | **Domain rating Y (yes) / PY (probably yes) / PN (probably no) / N (no) / NI (no information)** |
| Describe the sources of data and criteria for participant selection: Prospective cohort analysis in NESDA. However, excludes people who don't speak Dutch and comorbid psychiatric conditions. High comorbidity likely in severe populations therefore possible bias. Majority of NESDA sample = general population + primary care, however, this sample a higher proportion may be from secondary care (applicability concern). | |
| Were appropriate data sources used, e.g., cohort, randomized controlled trial, or nested case-control study data? | Y |
| Was an appropriate study design used? | Y |
| Did the in- and exclusions of study participants result in a representative dataset? | PN |
| DOMAIN 1 - OVERALL QUALITY CONCERN | HIGH |
| DOMAIN 1 - APPLICABILITY | HIGH |
| Domain 2 - Predictors | |
| List and describe predictors included in the final prediction model, how they were defined and assessed, and their timing of assessment: Baseline predictors (collected before outcome). Looks from supplement like all predictors were dichotomized – thresholds used were not clear e.g., what constitutes high/low income?-no evidence that consistent thresholds used. Many biological - likely available in a primary/secondary care setting, however maybe not all without conducting further blood tests ("probably yes"). Not explicitly stated whether pre-processing and definition/assessment was the same for all but assumed as no differences noted. | |
| Were predictors defined and assessed in a similar way for all participants? | PY / NI |
| Was any pre-processing of predictors similar for all participants? | NI |

| | |
|---|---|
| Were predictor assessments made without knowledge of outcome data? | PY |
| Are all predictors available at the time the model is intended to be used? | PY |
| DOMAIN 2 - OVERALL QUALITY CONCERN | LOW |
| DOMAIN 2 - APPLICABILITY | LOW |
| Domain 3 - Outcome | |
| Describe the outcome, how it was defined and determined, and the time interval between predictor assessment and outcome determination: Outcomes measured with validated scale. However, were self-reported so some bias (i.e., possible differences in reporting in those depressed vs non-depression – less objective) and predictors would be known to person reporting outcome (unblinded). Appropriate fit indices etc reported for trajectory modelling. Standard method of trajectory modelling, no applicability concern. 9 year follow up. | |
| Were outcomes defined and assessed appropriately? | PY |
| Was the outcome defined and determined in a similar way for all participants? | PN |
| Was the outcome determined without knowledge of predictor information? | PN |
| Was the time interval between predictor assessment and outcome determination appropriate? | Y |
| DOMAIN 3 - OVERALL QUALITY CONCERN | HIGH |
| DOMAIN 3 - APPLICABILITY | LOW |
| Domain 4 - Analysis | |
| Describe the numbers of participants, number of candidate predictors, number of outcome events. Describe how the prediction model was developed (e.g., with respect to modelling technique, predictor selection, and classification or risk group definition). Describe the performance measures of the prediction model, e.g., (re)calibration, discrimination, (re)classification, net benefit, and whether they were adjusted for optimism:<br>Overall sample, n=1693. Two models predicting probability of belonging to 3 trajectory groups each (continuous outcome = probability). Reported performance as MSE. No calibration or discrimination reported. EPV less relevant for continuous outcome but using binomial EPV calculation as a crude estimate, sample sizes likely small due to number of trajectory groups and high number of predictors (152). Cross-validation applied - helps DETECT overfitting but unclear if then adjusted? All categorical variables were recoded to dichotomous variables, coded as 0/1. Not clear what cut off was used. Continuous variables Z-transformed – normalise distribution. Predictors selected on availability in dataset rather than a univariable analysis. Only gives MSE as a performance measure. No mention of class imbalance – not relevant for continuous outcome?<br><br>Describe missing data on predictors and outcomes as well as methods used for handling these missing data:<br>Multiple imputation used and low missing data, also compared with CCAs in the supplement. 1.7% of the data were missing: 66/152 determinants had ≥1 missing value (range: 5.9%-15.8%. |  |
| Was model evaluation based on only apparent performance (i.e., training dataset only) avoided? | Y |
| Was there evidence that the sample size was reasonable? (development and evaluation samples) | N |
| Were continuous and categorical predictors handled appropriately? | PY/NI for some elements |
| Were participants with missing data handled appropriately? | PY |
| If methods to address class imbalance were used, was the model or the model predictions recalibrated? | NI (n/a) |

| | |
|---|---|
| If resampling methods were used to evaluate model performance, were all model development steps replicated in the resampling process? | NI |
| If data splitting was done to create training and test datasets, was there evidence that data leakage was avoided? | NI |
| Was the predictive performance of the model evaluated appropriately, e.g., calibration, discrimination, and net benefit? | N |
| Were methods used to address potential model overfitting? | PY |
| DOMAIN 4 - OVERALL QUALITY CONCERN | HIGH |

#### Dinga 2018

| | |
|---|---|
| **Publication reference** | **Dinga 2018** |
| **Model of interest (being evaluated)** | **Full model** |
| Domain 1 - Participants | **Domain rating Y (yes) / PY (probably yes) / PN (probably no) / N (no) / NI (no information)** |
| Describe the sources of data and criteria for participant selection: Prospective cohort analysis in NESDA. However, excludes people who don't speak Dutch and comorbid psychiatric conditions. High comorbidity likely in severe populations therefore possible bias. Majority of NESDA sample = general population + primary care, however, this sample a higher proportion may be from secondary care (applicability concern). | |
| Were appropriate data sources used, e.g., cohort, randomized controlled trial, or nested case-control study data? | Y |
| Was an appropriate study design used? | Y |
| Did the in- and exclusions of study participants result in a representative dataset? | PN |
| DOMAIN 1 - OVERALL QUALITY CONCERN | HIGH |
| DOMAIN 1 - APPLICABILITY | HIGH |
| Domain 2 - Predictors | |
| List and describe predictors included in the final prediction model, how they were defined and assessed, and their timing of assessment: Baseline predictors (collected before outcome). Many biological - likely available in a primary/secondary care setting, however maybe not all without conducting further blood tests ("probably yes"). Not explicitly stated whether pre-processing and definition/assessment was the same for all but assumed as no differences noted. | |
| Were predictors defined and assessed in a similar way for all participants? | PY / NI? |
| Was any pre-processing of predictors similar for all participants? | PY / NI? |
| Were predictor assessments made without knowledge of outcome data? | PY |

| | |
|---|---|
| Are all predictors available at the time the model is intended to be used? | PY |
| DOMAIN 2 - OVERALL QUALITY CONCERN | LOW |
| DOMAIN 2 - APPLICABILITY | LOW |
| Domain 3 - Outcome | |
| Describe the outcome, how it was defined and determined, and the time interval between predictor assessment and outcome determination: doi:10.1017/S0033291711002509 (for more information about trajectories). The LCI was assessed both at baseline and after 2 years of follow-up by a trained interviewer. Baseline LCI served to compute a measure of duration of depressive symptomatology. This methods means questions asked retrospectively (risk of bias). 24 data points - good number. Range of fit statistics given. No mention of outcome/predictor blinding. | |
| Were outcomes defined and assessed appropriately? | N |
| Was the outcome defined and determined in a similar way for all participants? | PY |
| Was the outcome determined without knowledge of predictor information? | PN |
| Was the time interval between predictor assessment and outcome determination appropriate? | PY |
| DOMAIN 3 - OVERALL QUALITY CONCERN | HIGH |
| DOMAIN 3 - APPLICABILITY | LOW |
| Domain 4 - Analysis | |
| Describe the numbers of participants, number of candidate predictors, number of outcome events. Describe how the prediction model was developed (e.g., with respect to modelling technique, predictor selection, and classification or risk group definition). Describe the performance measures of the prediction model, e.g., (re)calibration, discrimination, (re)classification, net benefit, and whether they were adjusted for optimism:<br><br>Logistic regression with elastic net penalty to reduce over fitting. Multinomial AND One-vs-rest (single binomial models; binary outcome). Best performing model had 81 predictors ("full model"). Risk of overfitting low for model with 5 predictors. EPV <5 for each trajectory prediction. No evidence of dichotomization of predictors or information about pre-processing of variables. "We focused on the biological variables that have shown to be related to depression or chronicity of depression in the previous cross-sectional studies" - likely initially a univariate selection process. Then stability selection process in model. Reported AUC (discrimination), and classification. No calibration. 10-fold cross validation.<br><br>Describe missing data on predictors and outcomes as well as methods used for handling these missing data: Imputation but from "similar individuals" (nearest neighbour? Knn?) rather than auxiliary variables in multiple imputation ("probably yes"). "The proportion of missing data was small; in total 25 out of 80 variables contained missing values. Of those variables, the median number of missing values per variable was 11 (1.4% of the sample). 23 out of 25 missing variables contained less than 7% of missing values with an exception of cortisol ROCg and cortisol ROCi with 38% of missing values." | |
| Was model evaluation based on only apparent performance (i.e., training dataset only) avoided? | Y |
| Was there evidence that the sample size was reasonable? (development and evaluation samples) | N |
| Were continuous and categorical predictors handled appropriately? | NI? |
| Were participants with missing data handled appropriately? | PY |

| | |
|---|---|
| If methods to address class imbalance were used, was the model or the model predictions recalibrated? | NI? |
| If resampling methods were used to evaluate model performance, were all model development steps replicated in the resampling process? | NI? |
| If data splitting was done to create training and test datasets, was there evidence that data leakage was avoided? | NI |
| Was the predictive performance of the model evaluated appropriately, e.g., calibration, discrimination, and net benefit? | N |
| Were methods used to address potential model overfitting? | Y |
| DOMAIN 4 - OVERALL QUALITY CONCERN | HIGH |

#### Kessler 2016

| **Publication reference** | **Kessler 2016** |
|---|---|
| **Model of interest (being evaluated)** | **ML model** |
| Domain 1 - Participants | **Domain rating Y (yes) / PY (probably yes) / PN (probably no) / N (no) / NI (no information)** |
| Describe the sources of data and criteria for participant selection: Two cross-sectional surveys; with a sub-sample reinterviewed (longitudinal prospective follow up in this sub-sample, probably yes - not as good as cohort). Target population - general US adult population. National household sample; not institutionalized. English speaking & household sample - may introduce some bias. Good response rate (83%). Weighting applied between surveys. Stratified, multistage area probability sample. 48 continuous United States. | |
| Were appropriate data sources used, e.g., cohort, randomized controlled trial, or nested case-control study data? | PY |
| Was an appropriate study design used? | PY |
| Did the in- and exclusions of study participants result in a representative dataset? | PY |
| DOMAIN 1 - OVERALL QUALITY CONCERN | LOW |
| DOMAIN 1 - APPLICABILITY | LOW |
| Domain 2 - Predictors | |
| List and describe predictors included in the final prediction model, how they were defined and assessed, and their timing of assessment: For baseline incident episode data - Fully structured interviews using CIDI, DSM-III-R criteria. "Blinded SCID21 clinical reappraisal interviews in a probability sub-sample found good concordance with DSM-III-R/CIDI diagnoses". Some predictors taken from survey 2 (i.e., not baseline) therefore outcome data known at this point & would not be available at time of prediction? Predictor types likely available in a clinical use setting (no applicability concerns). | |

| | |
|---|---|
| Were predictors defined and assessed in a similar way for all participants? | PY/NI? |
| Was any pre-processing of predictors similar for all participants? | PY/NI? |
| Were predictor assessments made without knowledge of outcome data? | N |
| Are all predictors available at the time the model is intended to be used? | PY |
| DOMAIN 2 - OVERALL QUALITY CONCERN | HIGH |
| DOMAIN 2 - APPLICABILITY | LOW |
| Domain 3 - Outcome | |
| Describe the outcome, how it was defined and determined, and the time interval between predictor assessment and outcome determination: Retrospective recall using life history calendar and computerized CIDI. Calculation of persistence/chronicity seems somewhat unusual. Unclear whether prespecified. Did not seem planned that they only used top 5-10% of persistence/chronic group - could introduce bias; cut off point not clinically determined. Outcomes self-reported so participants would be aware of own predictor info (not blinded). 10-12 year time frame good. Applicability concern as not standard/conventional trajectory formation method, although does meet our eligibility criteria for longitudinal pattern. | |
| Were outcomes defined and assessed appropriately? | N |
| Was the outcome defined and determined in a similar way for all participants? | PN |
| Was the outcome determined without knowledge of predictor information? | N |
| Was the time interval between predictor assessment and outcome determination appropriate? | Y |
| DOMAIN 3 - OVERALL QUALITY CONCERN | HIGH |
| DOMAIN 3 - APPLICABILITY | HIGH |
| Domain 4 - Analysis | |
| Describe the numbers of participants, number of candidate predictors, number of outcome events. Describe how the prediction model was developed (e.g., with respect to modelling technique, predictor selection, and classification or risk group definition). Describe the performance measures of the prediction model, e.g., (re)calibration, discrimination, (re)classification, net benefit, and whether they were adjusted for optimism:<br><br>Methods generally not detailed and often unclear. Used Ensemble regression trees to predict number of years/weeks; continuous outcome. Unable to locate sample size for persistent group. Overall sample n=1056. EPV likely <10 for chronic group. Talk about predictors in the WMH surveys (development study) but don't explicitly state whether predictors in this external validation dataset are the same. Selection based on the Van Loo 2014 paper which is called a "development" study but actually looks at univariate associations to justify model predictors. Unclear which predictors used. No calibration given. No CIs given with AUCs. Used 10 fold cross validation.<br><br>Describe missing data on predictors and outcomes as well as methods used for handling these missing data: Multiple imputation was applied to this dataset to generate 10 predicted scores on each missing variable to each Survey 1 respondent using SAS 9.2 proc mi. | |
| Was model evaluation based on only apparent performance (i.e., training dataset only) avoided? | Y |
| Was there evidence that the sample size was reasonable? (development and evaluation samples) | N |

| | |
|---|---|
| Were continuous and categorical predictors handled appropriately? | NI? |
| Were participants with missing data handled appropriately? | PY |
| If methods to address class imbalance were used, was the model or the model predictions recalibrated? | NI? |
| If resampling methods were used to evaluate model performance, were all model development steps replicated in the resampling process? | NI? |
| If data splitting was done to create training and test datasets, was there evidence that data leakage was avoided? | NI |
| Was the predictive performance of the model evaluated appropriately, e.g., calibration, discrimination, and net benefit? | N |
| Were methods used to address potential model overfitting? | Y |
| DOMAIN 4 - OVERALL QUALITY CONCERN | HIGH |

#### Wardenaar 2014

| | |
|---|---|
| **Publication reference** | **Wardenaar 2014** |
| **Model of interest (being evaluated)** | **Three cluster** |
| Domain 1 - Participants | **Domain rating Y (yes) / PY (probably yes) / PN (probably no) / N (no) / NI (no information)** |
| Describe the sources of data and criteria for participant selection: Retrospective cohort data. Representative, good global coverage in WMH surveys. At country level nationally representative. Target = general population. Weighted for selection and sociodemographic. Unclear which country/setting the model is intended to be applied to...? | |
| Were appropriate data sources used, e.g., cohort, randomized controlled trial, or nested case-control study data? | N |
| Was an appropriate study design used? | N |
| Did the in- and exclusions of study participants result in a representative dataset? | PY |
| DOMAIN 1 - OVERALL QUALITY CONCERN | HIGH |
| DOMAIN 1 - APPLICABILITY | LOW |
| Domain 2 - Predictors | |

| | |
|---|---|
| List and describe predictors included in the final prediction model, how they were defined and assessed, and their timing of assessment: retrospective; subject to recall bias which may be different between participants and means outcome/predictor info collected at same time. Low applicability or availability concerns; doesn't use any specialist measures. All related to incident episode or family history. | |
| Were predictors defined and assessed in a similar way for all participants? | PN |
| Was any pre-processing of predictors similar for all participants? | PY/NI? |
| Were predictor assessments made without knowledge of outcome data? | N |
| Are all predictors available at the time the model is intended to be used? | PY |
| DOMAIN 2 - OVERALL QUALITY CONCERN | HIGH |
| DOMAIN 2 - APPLICABILITY | LOW |
| Domain 3 - Outcome | |
| Describe the outcome, how it was defined and determined, and the time interval between predictor assessment and outcome determination: As with predictors - retrospectively collected; subject to recall bias. persistent depressive disorder in DSM-5 and DSM-5-TR is defined as a 2- year (or longer) period of depressed mood, most of the day, for more days than not. Authors only used top 5-10% of persistence/chronic group (more than 2 years, 10-15) - could introduce bias as this is only the top end of the sample; cut off point not clinically determined. Also they look at persistence and chronicity separately whereas DSM definition combines. Applicability concern as not standard/conventional trajectory formation method, although does meet our eligibility criteria for longitudinal pattern.. | |
| Were outcomes defined and assessed appropriately? | N |
| Was the outcome defined and determined in a similar way for all participants? | PN |
| Was the outcome determined without knowledge of predictor information? | N |
| Was the time interval between predictor assessment and outcome determination appropriate? | PY |
| DOMAIN 3 - OVERALL QUALITY CONCERN | HIGH |
| DOMAIN 3 - APPLICABILITY | HIGH |
| Domain 4 - Analysis | |
| Describe the numbers of participants, number of candidate predictors, number of outcome events. Describe how the prediction model was developed (e.g., with respect to modelling technique, predictor selection, and classification or risk group definition). Describe the performance measures of the prediction model, e.g., (re)calibration, discrimination, (re)classification, net benefit, and whether they were adjusted for optimism:<br><br>Methods generally not detailed and often unclear. 2869 subsample used to develop model; small number of predictors so EPV actually higher and less risk of overfitting associated with this. Not much information about data pre-processing. A brief mention of "dichotomous predictors" but not necessarily dichotomizing of originally continuous predictors. Selection based partially on the Van Loo 2014 paper which is called a "development" study but actually looks at univariate associations to justify model predictors. LASSO, Ridge and elastic net. 10-fold cross validation.<br><br>Describe missing data on predictors and outcomes as well as methods used for handling these missing data: Unclear – lack of information. | |
| Was model evaluation based on only apparent performance (i.e., training dataset only) avoided? | Y |

| | |
|---|---|
| Was there evidence that the sample size was reasonable? (development and evaluation samples) | PY |
| Were continuous and categorical predictors handled appropriately? | NI |
| Were participants with missing data handled appropriately? | NI |
| If methods to address class imbalance were used, was the model or the model predictions recalibrated? | NI |
| If resampling methods were used to evaluate model performance, were all model development steps replicated in the resampling process? | NI |
| If data splitting was done to create training and test datasets, was there evidence that data leakage was avoided? | NI |
| Was the predictive performance of the model evaluated appropriately, e.g., calibration, discrimination, and net benefit? | N |
| Were methods used to address potential model overfitting? | Y |
| DOMAIN 4 - OVERALL QUALITY CONCERN | HIGH |

#### Van Loo 2014

| **Publication reference** | **Van Loo 2014** |
|---|---|
| **Model of interest (being evaluated)** | **Three cluster** |
| Domain 1 - Participants | **Domain rating Y (yes) / PY (probably yes) / PN (probably no) / N (no) / NI (no information)** |
| Describe the sources of data and criteria for participant selection: Retrospective cohort data. Representative, good global coverage in WMH surveys. At country level nationally representative. Target = general population. Weighted for selection and sociodemographic. Unclear which country/setting the model is intended to be applied to...? | |
| Were appropriate data sources used, e.g., cohort, randomized controlled trial, or nested case-control study data? | N |
| Was an appropriate study design used? | N |
| Did the in- and exclusions of study participants result in a representative dataset? | PY |
| DOMAIN 1 - OVERALL QUALITY CONCERN | HIGH |
| DOMAIN 1 - APPLICABILITY | LOW |
| Domain 2 - Predictors | |

| | |
|---|---|
| List and describe predictors included in the final prediction model, how they were defined and assessed, and their timing of assessment: retrospective; subject to recall bias which may be different between participants and means outcome/predictor info collected at same time. Low applicability or availability concerns; doesn't use any specialist measures. All related to incident episode or family history. | |
| Were predictors defined and assessed in a similar way for all participants? | PN |
| Was any pre-processing of predictors similar for all participants? | PY/NI? |
| Were predictor assessments made without knowledge of outcome data? | N |
| Are all predictors available at the time the model is intended to be used? | PY |
| DOMAIN 2 - OVERALL QUALITY CONCERN | HIGH |
| DOMAIN 2 - APPLICABILITY | LOW |
| Domain 3 - Outcome | |
| Describe the outcome, how it was defined and determined, and the time interval between predictor assessment and outcome determination: As with predictors - retrospectively collected; subject to recall bias. persistent depressive disorder in DSM-5 and DSM-5-TR is defined as a 2- year (or longer) period of depressed mood, most of the day, for more days than not. Authors only used top 5-10% of persistence/chronic group (more than 2 years, 10-15) - could introduce bias as this is only the top end of the sample; cut off point not clinically determined. Also they look at persistence and chronicity separately whereas DSM definition combines. Applicability concern as not standard/conventional trajectory formation method, although does meet our eligibility criteria for longitudinal pattern.. | |
| Were outcomes defined and assessed appropriately? | N |
| Was the outcome defined and determined in a similar way for all participants? | PN |
| Was the outcome determined without knowledge of predictor information? | N |
| Was the time interval between predictor assessment and outcome determination appropriate? | PY |
| DOMAIN 3 - OVERALL QUALITY CONCERN | HIGH |
| DOMAIN 3 - APPLICABILITY | HIGH |
| Domain 4 - Analysis | |
| Describe the numbers of participants, number of candidate predictors, number of outcome events. Describe how the prediction model was developed (e.g., with respect to modelling technique, predictor selection, and classification or risk group definition). Describe the performance measures of the prediction model, e.g., (re)calibration, discrimination, (re)classification, net benefit, and whether they were adjusted for optimism:<br><br>Methods generally not detailed and often unclear. 2869 subsample used to develop model; small number of predictors so EPV actually higher and less risk of overfitting associated with this. Not much information about data pre-processing. A brief mention of "dichotomous predictors" but not necessarily dichotomizing of originally continuous predictors. Selection based partially on the Van Loo 2014 paper which is called a "development" study but actually looks at univariate associations to justify model predictors. LASSO, Ridge and elastic net. 10-fold cross validation.<br><br>Describe missing data on predictors and outcomes as well as methods used for handling these missing data: Unclear – lack of information. | |
| Was model evaluation based on only apparent performance (i.e., training dataset only) avoided? | Y |

| | |
|---|---|
| Was there evidence that the sample size was reasonable? (development and evaluation samples) | PY |
| Were continuous and categorical predictors handled appropriately? | NI |
| Were participants with missing data handled appropriately? | NI |
| If methods to address class imbalance were used, was the model or the model predictions recalibrated? | NI |
| If resampling methods were used to evaluate model performance, were all model development steps replicated in the resampling process? | NI |
| If data splitting was done to create training and test datasets, was there evidence that data leakage was avoided? | NI |
| Was the predictive performance of the model evaluated appropriately, e.g., calibration, discrimination, and net benefit? | N |
| Were methods used to address potential model overfitting? | Y |
| DOMAIN 4 - OVERALL QUALITY CONCERN | HIGH |

#### Gorham 2022

| **Publication reference** | **Gorham 2022** |
|---|---|
| **Model of interest (being evaluated)** | **MFQ+FH+CASE** |
| Domain 1 - Participants | **Domain rating Y (yes) / PY (probably yes) / PN (probably no) / N (no) / NI (no information)** |
| Describe the sources of data and criteria for participant selection: longitudinal cohort. However very small sample (92) and several exclusions which may introduce bias such as: unwilling to undergo psychiatric interviews, cannot speak English, does not have a primary care clinician in the community, other psychiatric or neurodevelopmental conditions, IQ <70, depressive symptoms that are due to effects of drugs of abuse or a neurological condition; a diagnosis of alcohol or other substance use disorders; current active suicidal ideation; repeated self-harm in the context of interpersonal conflict; having an immediate family member who works at the NIMH; and a serious medical condition such as epilepsy or heart disease. | |
| Were appropriate data sources used, e.g., cohort, randomized controlled trial, or nested case-control study data? | Y |
| Was an appropriate study design used? | N |
| Did the in- and exclusions of study participants result in a representative dataset? | N |
| DOMAIN 1 - OVERALL QUALITY CONCERN | HIGH |
| DOMAIN 1 - APPLICABILITY | LOW |

| | |
|---|---|
| Domain 2 - Predictors | |
| List and describe predictors included in the final prediction model, how they were defined and assessed, and their timing of assessment: all measured at baseline other than ACE; which was measured at 1 year follow up. Would have happened before outcome but measured after with knowledge of outcome. "Since the CASE was not collected at baseline, we did not use it in these models" – however best performing model did use CASE? no specialist measures used, and few of them - likely available and no applicability concerns | |
| Were predictors defined and assessed in a similar way for all participants? | PY |
| Was any pre-processing of predictors similar for all participants? | PY/NI? |
| Were predictor assessments made without knowledge of outcome data? | N |
| Are all predictors available at the time the model is intended to be used? | PY |
| DOMAIN 2 - OVERALL QUALITY CONCERN | HIGH |
| DOMAIN 2 - APPLICABILITY | LOW |
| Domain 3 - Outcome | |
| Describe the outcome, how it was defined and determined, and the time interval between predictor assessment and outcome determination: Kiddy-SADS. All cases were discussed between two senior child psychiatrists. Not all participants had the same number of repeated measures. Pre-registered analysis: "number of weeks spent in a depressive episode from the baseline visit to the one-year follow-up"... not conventional trajectory method. Unclear if assessors blinded to depression diagnosis @ baseline? Therefore weeks of depression assessment may be influenced by this? 1 year follow up possibly too short | |
| Were outcomes defined and assessed appropriately? | N |
| Was the outcome defined and determined in a similar way for all participants? | PY |
| Was the outcome determined without knowledge of predictor information? | NI |
| Was the time interval between predictor assessment and outcome determination appropriate? | PN |
| DOMAIN 3 - OVERALL QUALITY CONCERN | HIGH |
| DOMAIN 3 - APPLICABILITY | HIGH |
| Domain 4 - Analysis | |
| Describe the numbers of participants, number of candidate predictors, number of outcome events. Describe how the prediction model was developed (e.g., with respect to modelling technique, predictor selection, and classification or risk group definition). Describe the performance measures of the prediction model, e.g., (re)calibration, discrimination, (re)classification, net benefit, and whether they were adjusted for optimism:<br><br>Methods not detailed. Very small sample (92). Only RMSE given as performance. Used bootstrapping and cross validation.<br><br><br>Describe missing data on predictors and outcomes as well as methods used for handling<br>these missing data: Unclear – lack of information. no mention of imputation and mentions excluding CASE due to high level of missing data and several tables in supplement have "requirements for inclusion" that mention availability of data. | |
| Was model evaluation based on only apparent performance (i.e., training dataset only) avoided? | Y |

| | |
|---|---|
| Was there evidence that the sample size was reasonable? (development and evaluation samples) | N |
| Were continuous and categorical predictors handled appropriately? | NI |
| Were participants with missing data handled appropriately? | N |
| If methods to address class imbalance were used, was the model or the model predictions recalibrated? | NI |
| If resampling methods were used to evaluate model performance, were all model development steps replicated in the resampling process? | NI |
| If data splitting was done to create training and test datasets, was there evidence that data leakage was avoided? | NI |
| Was the predictive performance of the model evaluated appropriately, e.g., calibration, discrimination, and net benefit? | N |
| Were methods used to address potential model overfitting? | N |
| DOMAIN 4 - OVERALL QUALITY CONCERN | HIGH |

#### Schultebraucks 2021

| | |
|---|---|
| Publication reference | Schultebraucks 2021 |
| Model of interest (being evaluated) | Neural network |
| Domain 1 - Participants | **Domain rating Y (yes) / PY (probably yes) / PN (probably no) / N (no) / NI (no information)** |
| Describe the sources of data and criteria for participant selection: prospective longitudinal cohort. European ancestry only – uses genetic data. Higher proportion of women. Older adults with major life stressor – applicability concern? | |
| Were appropriate data sources used, e.g., cohort, randomized controlled trial, or nested case-control study data? | Y |
| Was an appropriate study design used? | Y |
| Did the in- and exclusions of study participants result in a representative dataset? | N |
| DOMAIN 1 - OVERALL QUALITY CONCERN | HIGH |
| DOMAIN 1 - APPLICABILITY | HIGH |
| Domain 2 - Predictors | |

| | |
|---|---|
| List and describe predictors included in the final prediction model, how they were defined and assessed, and their timing of assessment: genotyping process detailed in supplement. Applicability issue with availability of genome data in the RW (count this as applicability concern). However, temporally genetic data always precedes outcome and cannot be influenced by bias as subjective. | |
| Were predictors defined and assessed in a similar way for all participants? | Y |
| Was any pre-processing of predictors similar for all participants? | NI? |
| Were predictor assessments made without knowledge of outcome data? | Y |
| Are all predictors available at the time the model is intended to be used? | PN |
| DOMAIN 2 - OVERALL QUALITY CONCERN | LOW |
| DOMAIN 2 - APPLICABILITY | HIGH |
| Domain 3 - Outcome | |
| Describe the outcome, how it was defined and determined, and the time interval between predictor assessment and outcome determination: CES-D = appropriate validated measure (yes). However, was self-reported (not standardized/less objective so may differ). PGS used for depression phenotype… does this count as predictor and outcome overlap?? Especially for "pre-existing" trajectory. Participants will be blinded to genome but not phenotype. 2 year follow up. | |
| Were outcomes defined and assessed appropriately? | Y |
| Was the outcome defined and determined in a similar way for all participants? | PN |
| Was the outcome determined without knowledge of predictor information? | PY |
| Was the time interval between predictor assessment and outcome determination appropriate? | PY |
| DOMAIN 3 - OVERALL QUALITY CONCERN | HIGH |
| DOMAIN 3 - APPLICABILITY | LOW |
| Domain 4 - Analysis | |
| Describe the numbers of participants, number of candidate predictors, number of outcome events. Describe how the prediction model was developed (e.g., with respect to modelling technique, predictor selection, and classification or risk group definition). Describe the performance measures of the prediction model, e.g., (re)calibration, discrimination, (re)classification, net benefit, and whether they were adjusted for optimism:<br><br>/AUC curves and F1. Discrimination only? No calibration? Multinomial model – EPV of 1.81.<br>Categorical variables were dummy coded into binary values. Continuous variables were normalized to the range of [0;1]<br>Cross validation with loops to adjust for overfitting - described in supplement<br><br>Describe missing data on predictors and outcomes as well as methods used for handling<br>these missing data: Appears to be complete case analysis - only included participants with trajectory information and single stressor information available. “variables with more than 35% missingness were removed”. ROC | |
| Was model evaluation based on only apparent performance (i.e., training dataset only) avoided? | Y |
| Was there evidence that the sample size was reasonable? (development and evaluation samples) | N |
| Were continuous and categorical predictors handled appropriately? | PY |

| | |
|---|---|
| Were participants with missing data handled appropriately? | PN |
| If methods to address class imbalance were used, was the model or the model predictions recalibrated? | NI |
| If resampling methods were used to evaluate model performance, were all model development steps replicated in the resampling process? | NI |
| If data splitting was done to create training and test datasets, was there evidence that data leakage was avoided? | NI |
| Was the predictive performance of the model evaluated appropriately, e.g., calibration, discrimination, and net benefit? | N |
| Were methods used to address potential model overfitting? | N |
| DOMAIN 4 - OVERALL QUALITY CONCERN | HIGH |

#### Teutenberg 2025

| | |
|---|---|
| Publication reference | Teutenberg 2025 |
| Model of interest (being evaluated) | Explorative model |
| Domain 1 - Participants | **Domain rating Y (yes) / PY (probably yes) / PN (probably no) / N (no) / NI (no information)** |
| Describe the sources of data and criteria for participant selection: Excluded: any life-time substance dependence disorder, had a history of severe neurological or medical disorders, did not have a West-European ancestry. This may introduce bias. Prospective cohort – paper suggests includes population based, primary and secondary care. However, other papers imply secondary. Not much information on the cohort (applicability issue). | |
| Were appropriate data sources used, e.g., cohort, randomized controlled trial, or nested case-control study data? | Y |
| Was an appropriate study design used? | Y |
| Did the in- and exclusions of study participants result in a representative dataset? | N |
| DOMAIN 1 - OVERALL QUALITY CONCERN | HIGH |
| DOMAIN 1 - APPLICABILITY | HIGH |
| Domain 2 - Predictors | |
| List and describe predictors included in the final prediction model, how they were defined and assessed, and their timing of assessment: Predictors measured at baseline. Not highly specialised predictors e.g., genome or biomarkers; could all probably be obtained via patient report. | |
| Were predictors defined and assessed in a similar way for all participants? | PY? |

| | |
|---|---|
| Was any pre-processing of predictors similar for all participants? | NI? |
| Were predictor assessments made without knowledge of outcome data? | Y |
| Are all predictors available at the time the model is intended to be used? | PY |
| DOMAIN 2 - OVERALL QUALITY CONCERN | LOW |
| DOMAIN 2 - APPLICABILITY | LOW |
| Domain 3 - Outcome | |
| Describe the outcome, how it was defined and determined, and the time interval between predictor assessment and outcome determination: Reported retrospectively using life charting method. Only over 2 year period so better than a longer period (i.e., less opportunity for recall bias) but still bias possible. 2 years may be too short. Trained interviewers (objective), no mention of blinding. | |
| Were outcomes defined and assessed appropriately? | N |
| Was the outcome defined and determined in a similar way for all participants? | PN |
| Was the outcome determined without knowledge of predictor information? | NI? |
| Was the time interval between predictor assessment and outcome determination appropriate? | PY |
| DOMAIN 3 - OVERALL QUALITY CONCERN | HIGH |
| DOMAIN 3 - APPLICABILITY | LOW |
| Domain 4 - Analysis | |
| Describe the numbers of participants, number of candidate predictors, number of outcome events. Describe how the prediction model was developed (e.g., with respect to modelling technique, predictor selection, and classification or risk group definition). Describe the performance measures of the prediction model, e.g., (re)calibration, discrimination, (re)classification, net benefit, and whether they were adjusted for optimism:<br><br>Data pre-processing not clear. Supp does mention RF can handle both categorical and continuous predictors. One-vs-rest binomial models. EPV <4 for all. Nested cross-validation. AUC. No calibration given. Does compare results across validation and test samples. “To handle class imbalance in predictions, we implemented cross-validation using stratification, ensuring that the proportion of each class remained consistent across all folds. As class imbalance was especially occurring in the prediction of MDD trajectories, we further utilized balanced RFCs in all models predicting trajectories. Thus, the classifier operated by creating a balanced bootstrap sample by randomly under-sampling the majority class for each tree build, ensuring that each tree was trained on a balanced dataset.”<br><br>Describe missing data on predictors and outcomes as well as methods used for handling these missing data: Missing data for trajectories - n=14 exclusions (CCA). Relatively small sample of missing so minimal bias. In prediction, all missing data <5% and KNN imputation used. | |
| Was model evaluation based on only apparent performance (i.e., training dataset only) avoided? | Y |
| Was there evidence that the sample size was reasonable? (development and evaluation samples) | N |
| Were continuous and categorical predictors handled appropriately? | NI |
| Were participants with missing data handled appropriately? | PY |

| | |
|---|---|
| If methods to address class imbalance were used, was the model or the model predictions recalibrated? | Y |
| If resampling methods were used to evaluate model performance, were all model development steps replicated in the resampling process? | PY? |
| If data splitting was done to create training and test datasets, was there evidence that data leakage was avoided? | NI |
| Was the predictive performance of the model evaluated appropriately, e.g., calibration, discrimination, and net benefit? | N |
| Were methods used to address potential model overfitting? | Y |
| DOMAIN 4 - OVERALL QUALITY CONCERN | HIGH |

### Feature/predictor lists for main models

***Lists of features included in each of the main models and top 10-15 features for each study.***

#### Xiang 2022

24 features included in main GBM model, **only the complete list of candidates and the top 11 are listed in paper.**

Top 10 features:

1. Total sleep disturbance score
2. Total problems of parent adult self-report syndrome
3. Number of financial adversities in family in past 12 months
4. Average correlation between ventral attention network and ASEG ROI left-caudate
5. Average correlation between dorsal attention network and ASEG ROI left-caudate
6. Score of school disengagement subscale
7. Average correlation between salience network and ASEG ROI left-accumbens-area
8. Residential history derived – cross residential density
9. Score of school involvement subscale
10. Score of parental monitoring and supervision

#### Wardenaar 2021

##### Baseline predictor list (n=152)

1. Gender
2. Partner
3. Employment
4. Father died before age 16
5. Mother died before age 16
6. Having children
7. Smoking
8. Drug use
9. Sleeping problems (insomnia rating scale>9)
10. Any somatic disease
11. Any treatment for somatic disease
12. Pain (CPG=3 or 4)
13. Separation before age 16
14. Parents divorced
15. Death of a parent before age 16
16. Use of hypertensive drugs
17. Hypertension
18. High waist circumference
19. High triglycerides
20. Low HDL cholesterol
21. Hypertension
22. High blood glucose
23. Metabolic syndrome
24. non-NL country of birth
25. Low income
26. High income
27. Money problems
28. Alcohol problems

29. Any childhood life event
30. Childhood life event index>1
31. Any childhood trauma
32. Childhood trauma index=4
33. Childhood trauma index>=3
34. Childhood trauma index>=2
35. 1-M minor depression
36. 1-M Dysthymia
37. 1-M Major Depressive Disorder
38. 6-M Dysthymia
39. 6-M Major Depressive Disorder
40. Any recurrence
41. Previous episodes: 1-3
42. Previous episodes: 4-6
43. Previous episodes: 7-10
44. Previous episodes: >10
45. Any 1-M depression
46. Any 6-M depression
47. 1-M Social Anxiety Disorder
48. 1-M Panic Disorder w/ Agoraphobia
49. 1-M Panic Disorder
50. 1-M Agoraphobia
51. 1-M Generalized Anxiety Disorder
52. 2 or more 1-M anxiety disorders
53. 3 or more 1-M anxiety disorders
54. 6-M Social Anxiety Disorder
55. 6-M Panic Disorder w/ Agoraphobia
56. 6-M Panic Disorder
57. 6-M Agoraphobia
58. 6-M Generalized Anxiety Disorder
59. 2 or more 6-M anxiety disorders
60. 3 or more 6-M anxiety disorders
61. Any 1-M anxiety disorder

62. Any 6-M anxiety disorder
63. Any 1-M comorbidity (anx + dep)
64. Any 6-M comorbidity (anx + dep)
65. two comorbid 1-M disorders
66. three comorbid 1-M disorders
67. four or more comorbid 1-M disorders
68. two comorbid 6-M disorders
69. three comorbid 6-M disorders
70. four or more comorbid 6-M disorders
71. Latest recency of any disorder is in the past month
72. Ever a suicide attempt
73. Any suicidal thoughts in last week
74. Any threatening life event (LTE)
75. two or more LTEs
76. three or more LTEs
77. Use of homecare
78. Use of alternative healthcare
79. Visit a self-help group
80. Visit a hospital
81. Visit a physician
82. Visit a medical specialist
83. Visit an occupational physician
84. Visit a psychologist
85. Visit a mental healthcare (MH) institution
86. Substance-use treatment
87. Visit an idependent psychatrist
88. Visit a physiotherapist
89. Use of tricyclic antidepressants
90. Use of SSRIs
91. Use of other antidepressants
92. Use of benzodiazepines
93. Use of any psychotropic medication
94. Use of any psycholepitic medication

95. Use of antipsychotics
96. Use of any antidepressants
97. Having a kidney disease
98. Age
99. Years of education
100. Insomnia rating scale totals score
101. IPAQ total score
102. MASQ-D30 General Distress
103. MASQ-D30 Anhedonic Depression
104. MASQ-D30 Anxious Arousal
105. MDQ score
106. De Jong Gierveld Loneliness score
107. De Jong Gierveld emotional Loneliness score
108. De Jong Gierveld social Loneliness score
109. 4DSQ Distress scale
110. Mastery scale
111. Daily hassles: daily pressure
112. Daily hassles: daily rejection
113. NEO-FFI Neuroticism
114. NEO-FFI Extraversion
115. NEO-FFI openness
116. NEO-FFI Agreeableness
117. NEO-FFI Conscientiousness
118. CPG pain intensity score
119. CPG pain disability score
120. Youngest AOO across disorders
121. 4DSQ somatization
122. WHODAS Total score
123. WHODAS Cognition score
124. WHODAS Mobility score
125. WHODAS Self-care score
126. WHODAS getting along with other people score
127. WHODAS Life activities score

128. WHODAS Participation score
129. Heartrate
130. Root mean square of successive differences (RMSSD)
131. Standard deviation of NN intervals (SDNN)
132. Respiratory Sinus Arrythmia (RSA)
133. Respiratory rate (RR)
134. Preejection period (PEP)
135. Gamma-GT level
136. Aspartate transaminase (ASAT-GOT) level
137. Alanine Aminotransferase (ALAT-GPT) level
138. Glucose level
139. Triglycerides level
140. HDL cholesterol level
141. LDL cholesterol level
142. Haemoglobin (HB)
143. Haemetocrite (HT)
144. Erythrocites (Erys)
145. C-reactive proteine (CRP)
146. Interleukine-6 (il6)
147. Tumor-necrosis factor alpha (TNF-alpha)
148. Brain derived neurotrophic factor (BDNF)
149. Tryptophan (Tryp)
150. Kynurenine (Kyn)
151. Creatinine
152. Glomerular filtration rate (GFR)

**The following emerged in list of top 15 predictors across trajectories (note that there are more than 15 because each trajectory group had its own set of important predictors but there is overlap between trajectories):**

1. Having children
2. Any somatic disease
3. Metabolic syndrome

4. non-NL country of birth
5. Low income
6. Childhood trauma index>=2
7. 1-M minor depression
8. 1-M Dysthymia
9. Previous episodes: 4-6
10. Previous episodes: >10
11. Any 1-M depression
12. 1-M Panic Disorder
13. 1-M Agoraphobia
14. 2 or more 6-M anxiety disorders
15. 3 or more 6-M anxiety disorders
16. three comorbid 1-M disorders
17. Ever a suicide attempt
18. Visit a physician
19. Visit a medical specialist
20. Visit an occupational physician
21. Visit a psychologist
22. Visit a mental healthcare (MH) institution
23. Substance-use treatment
24. Visit a physiotherapist
25. Use of any antidepressants
26. Age
27. Years of education
28. MASQ-D30 General Distress
29. MASQ-D30 Anhedonic Depression
30. MASQ-D30 Anxious Arousal
31. MDQ score
32. De Jong Gierveld Loneliness score
33. De Jong Gierveld emotional Loneliness score
34. De Jong Gierveld social Loneliness score
35. 4DSQ Distress scale
36. Mastery scale

37. Daily hassles: daily pressure
38. NEO-FFI Neuroticism
39. NEO-FFI Extraversion
40. NEO-FFI openness
41. Youngest AOO across disorders
42. 4DSQ somatization
43. WHODAS getting along with other people score
44. WHODAS Life activities score
45. WHODAS Participation score
46. Heartrate
47. Standard deviation of NN intervals (SDNN)
48. Respiratory rate (RR)
49. Preejection period (PEP)
50. Alanine Aminotransferase (ALAT-GPT) level
51. Haemetocrite (HT)
52. C-reactive proteine (CRP)
53. Interleukine-6 (il6)
54. Glomerular filtration rate (GFR)

#### Dinga 2018

**Authors state that full model has 81 predictors, the following are listed in the supplement (does not add up to 81):**

1. Summary score of the Inventory of Depressive Symptomatology questionnaire (IDS).
2. 9 PREDICTORS - MDD and dysthymia related measures were derived from the CIDI: presence of each of the 9 individual MDD DSM-IV criteria (yes/no score for each of the 9 MDD criteria)

3. 3 PREDICTORS: severity of MDD (yes/no: mild, moderate, severe),
4. 4 PREDICTORS: recency of MDD (4 yes/no indicators for presence of MDD in past 1 month, 6 months, 12 months and life-time)
5. age of onset of MDD
6. 4 PREDICTORS: presence and recency of dysthymia (4 yes/no indicators for presence of dysthymia in past 1 month, 6 months, 12 months and life-time),
7. recurrence of MDD (yes/no first episode)
8. number of prior MDD episodes
9. anxiety severity measured by the Beck Anxiety Inventory (BAI)
10. 5 PREDICTORS: the presence or absence of each of the following DSM-IV anxiety disorders assessed by the CIDI: social phobia, agoraphobia, panic disorder, panic agoraphobia and general anxiety disorder
11. 4 PREDICTORS: recency of each of the anxiety disorders (yes/no presence of each disorder in 1, 6 and 12 months and lifetime)
12. total number of current anxiety diagnoses.
13. Childhood trauma (before the age of 16) assessed using a childhood trauma interview. The total childhood trauma score was calculated as the sum (0-8) of number and frequency of traumatic events scored 0-2 for 4 domains: emotional neglect, psychological abuse, physical abuse and sexual abuse
14. Family history was evaluated using the family tree method. Persons with a first-degree family member with depression or anxiety were considered to have a positive family history.
15. Waist circumference was measured using a measuring tape at the central point between lowest front rib and highest point of pelvis.
16. Vitamin D (fasting blood samples, collected in the morning of baseline interview and kept frozen).
17. CRP (fasting blood samples, collected in the morning of baseline interview and kept frozen).
18. IL6 (fasting blood samples, collected in the morning of baseline interview and kept frozen).
19. TNF-alpha (fasting blood samples, collected in the morning of baseline interview and kept frozen).
20. Cortisol levels were assessed from saliva samples using Salivettes
21. BMI was calculated as measured weight in kilograms divided by squared height in meters.
22. hand-grip strength (Jamar hand-held dynamo-meter, during a sitting, straight backed)
23. lung function - peak expiatory flow using a mini Wright peak flow meter
24. The total number of chronic diseases was established based on a self-report questionnaire, and included lung disease, diabetes, cardiovascular disease, cancer, osteoarthritis, intestinal disorder, liver disease, epilepsy and thyroid gland diseases for which a respondent received treatment and/or medical attention.
25. Heart rate (VU-AMS)
26. heart rate variability (VU-AMS)
27. ECG was assessed using a six-electrode configuration.

**The following top 9 predictors are listed:**

1. IDS score
2. Conscientiousness
3. Extraversion
4. Suicidality
5. Dysthymia lifetime
6. Dysthymia 12m
7. Dysthymia 6m
8. Dysthymia 1m
9. Dysthymia

#### Kessler 2016

*This model was based on Van loo 2014 and Wardenaar 2014 (below). It is unclear which predictors were available in the validation dataset that was used and how these were measured. It appears that there are differences between the predictors available in the development set and this validation study e.g., the list of comorbid conditions differs. The following list have been extracted as a best guess of likely predictors included in models.*

**Following 23 predictors in logistic regression model. Authors state that "ML model contained fewer predictors, 9-13" but clear which.**

1. Age of onset
2. Whether first ever depressive episode brought on by a stressful experience or spontaneous
3. 9 DSM-IV CIDI Criterion A-D symptoms of MDE for the index episode
4. Binary variable for respondent Family History Research Diagnostic Criteria Interview reports for whether respondents' parents had a history of major depression.
5. Additional items from the DSM-IV CIDI (additional to MDD) which includes sections for the following 11 disorders (**6 anxiety disorders and 5 externalizing disorders)**, each section has questions about age of onset, duration and severity:
    a) GAD
    b) Panic disorder

    c) PTSD
    d) Specific phobia
    e) Agoraphobia
    f) Social phobia
    g) Alcohol abuse with or without dependence
    h) Drug abuse with or without dependence
    i) Alcohol dependence
    j) Drug dependence
    k) Mania/hypomania

#### Van Loo 2014

**List of predictors for this paper is not clear. The following predictors have been extracted as a best guess from the paper. It is unclear how many individual predictor items were generated from each concept in this list.**

**Estimated total at least 29:**

1. Age of onset
2. Whether first ever depressive episode brought on by a stressful experience or spontaneous
3. All DSM-IV CIDI Criterion A-D symptoms of MDE for the index episode (including separate questions about):
    a) weight loss
    b) weight gain
    c) insomnia
    d) hypersomnia
    e) psychomotor agitation
    f) retardation
    g) thoughts of death
    h) suicide ideation
    i) suicide plans
    j) suicide gestures-attempts
4. ICD-10 severity specifiers
    a) Mild

   b) Moderate
   c) Severe
5. questions to operationalize diagnostic hierarchy rule exclusions *(NB: unclear what this means or how many questions)*
6. questions about symptoms during the index episode that might be markers of:
   a) Dysthymia (inability to cope; social withdrawal)
   b) Mixed episodes (sleep less than usual and still not feeling too tired, racing thoughts)
   c) Anxious depression (feeling irritable, nervous-anxious-worried, sudden attacks of intense fear or panic)
7. Binary variable for respondent Family History Research Diagnostic Criteria Interview reports for whether respondents' parents had a history of major depression.
8. Eight nested categories about respondents AOO
9. AAI – AOO onset categories (unclear how many)

#### Wardenaar 2014

**This paper was based on Van Loo 2014 but included an expanded set of predictors relating to prior lifetime comorbidities. The list of predictors for this paper is still not clear. The following predictors have been extracted as a best guess from the paper. It is unclear how many individual predictor items were generated from each concept in this list.**

**Estimated total at least 41:**

6. Age of onset
7. Whether first ever depressive episode brought on by a stressful experience or spontaneous
8. All DSM-IV CIDI Criterion A-D symptoms of MDE for the index episode (including separate questions about):
   a) weight loss
   b) weight gain
   c) insomnia
   d) hypersomnia
   e) psychomotor agitation
   f) retardation
   g) thoughts of death
   h) suicide ideation
   i) suicide plans
   j) suicide gestures-attempts

9. ICD-10 severity specifiers
    a) Mild
    b) Moderate
    c) Severe
10. questions to operationalize diagnostic hierarchy rule exclusions *(NB: unclear what this means or how many questions)*
11. questions about symptoms during the index episode that might be markers of:
    a) Dysthymia (inability to cope; social withdrawal)
    b) Mixed episodes (sleep less than usual and still not feeling too tired, racing thoughts)
    c) Anxious depression (feeling irritable, nervous-anxious-worried, sudden attacks of intense fear or panic)
12. Binary variable for respondent Family History Research Diagnostic Criteria Interview reports for whether respondents' parents had a history of major depression.
13. Additional items from the DSM-IV CIDI (additional to MDD) which includes sections for the following **14 disorders**, each section has questions about age of onset, duration and severity:
    a) GAD
    b) Panic disorder
    c) PTSD
    d) Social phobia
    e) Alcohol/drug abuse with or without dependence
    f) Substance dependence
    g) Mania/hypomania
    h) Psychosis
    i) OCD
    j) Suicidality
    k) Separation anxiety
    l) Specific phobia
    m) Agoraphobia
    n) ADHD
    o) Intermitted explosive disorder
    p) Oppositional-defiant disorder
    q) Conduct disorder
14. In addition to considering the list above in point 17 alone, also aggregated combinations of these. These included the **following 8:**
    a) 1+ distress disorders
    b) 2+ distress disorders

c) 1+ fear disorders
d) 2+ fear disorders
e) 3+ fear disorders
f) Child adolescent onset
g) Adult onset
h) Child-adolescent onset less than or equal to age 18

#### Gorham 2022

**10 items in the MFQ+FH+CASE model which was best performing:**

1. Baseline MFQ
2. Family history of depression
3. Antidepressants at Baseline
4. Other Meds at Baseline
5. Antidepressants at follow up
6. Other Meds at follow up
7. Inpatient
8. Sex
9. Age
10. Pandemic

#### Teutenberg 2025

**20 variables were included as predictors in “explorative” model (best performing) – only 18 listed but possible that authors forgot to list introversion and neuroticism under NEO FFI:**

1. Prior hospitalisations
2. Familial risk (presence/absence of any affective disorder diagnosis or treatment in 1$^{st}$ degree relative)
3. Hamilton Anxiety Scale
4. Young Mania Rating scale
5. Scale for the Assessment of Negative Symptoms

6. Scale for the Assessment of Positive Symptoms
7. NEO FFI: extraversion
8. NEO FFI: openness
9. NEO FFI: agreeableness
10. NEO FFI: conscientiousness
11. Current social support questionnaire
12. Childhood maternal and parental care (parental bonding instrument)
13. RS-25 resilience questionnaire
14. Age
15. Sex
16. Years of education
17. Urbanicity (Lederbogen urban upbringing score)
18. Stressful life events in past 6 months

#### Schultebraucks 2021

**Polygenic risk scores for the following 21 phenotypes. Nearly all other than smoking behaviour, OCD, bipolar disorder and cortisol were important features for predicting at least one trajectory group:**

1. Major depressive disorder
2. Schizophrenia
3. Posttraumatic stress disorder
4. Attention-deficit/hyperactivity disorder
5. Obsessive-compulsive disorder
6. Bipolar disorder
7. Neuroticism
8. Depressive symptoms
9. Anxiety symptoms continuous
10. Well-being
11. Extraversion
12. Cognitive function
13. Educational attainment
14. Cortisol
15. Smoking behaviour

16. Waist circumference
17. Body fat distribution
18. Body mass index
19. High-density lipoprotein cholesterol
20. Low-density lipoprotein cholesterol
21. Total cholesterol

90. Accuracy of general practitioner's prognosis of the 1-year course of depression and generalised anxiety | The British Journal of Psychiatry | Cambridge Core. https://www.cambridge.org/core/journals/the-british-journal-of-psychiatry/article/accuracy-of-general-practitioners-prognosis-of-the-1year-course-of-depression-and-generalised-anxiety/503BC38DA6C588E50A0F4AAA625FA316.

123. The World Mental Health Survey Initiative. https://www.hcp.med.harvard.edu/wmh/national_sample.php?utm_source=chatgpt.com.

124. Fisher, G. G. & Ryan, L. H. Overview of the Health and Retirement Study and Introduction to the Special Issue. *Work Aging Retire* 4, 1–9 (2018) doi:10.1093/workar/wax032.
